# Trends in risk factors and symptoms associated with SARS-CoV-2 and Rhinovirus test positivity in King County, Washington: A Test-Negative Design Study of the Greater Seattle Coronavirus Assessment Network

**DOI:** 10.1101/2022.08.12.22278203

**Authors:** Chelsea L. Hansen, Amanda Perofsky, Roy Burstein, Michael Famulare, Shanda Boyle, Robin Prentice, Cooper Marshall, Benjamin JJ McCormick, David Reinhart, Ben Capodanno, Melissa Truong, Kristen Schwabe-Fry, Kayla Kuchta, Brian Pfau, Zack Acker, Jover Lee, Thomas R. Sibley, Evan McDermot, Leslie Rodriguez-Salas, Jeremy Stone, Luis Gamboa, Peter D. Han, Jeffery S. Duchin, Alpana Waghmare, Janet A. Englund, Jay Shendure, Trevor Bedford, Helen Y. Chu, Lea M. Starita, Cécile Viboud

## Abstract

**Importance:** Few US studies have reexamined risk factors for SARS-CoV-2 positivity in the context of widespread vaccination and new variants or considered risk factors for co-circulating endemic viruses, such as rhinovirus.

**Objective:** To understand how risk factors and symptoms associated with SARS-CoV-2 test positivity changed over the course of the pandemic and to compare these to the factors associated with rhinovirus test positivity.

**Design:** This test-negative design study used multivariable logistic regression to assess associations between SARS-CoV-2 and rhinovirus test positivity and self-reported demographic and symptom variables over a 22-month period.

**Setting:** King County, Washington, June 2020-April 2022

**Participants:** 23,278 symptomatic individuals of all ages enrolled in a cross-sectional community surveillance study.

**Exposures:** Self-reported data for 15 demographic and health behavior variables and 16 symptoms.

**Main Outcome(s) and Measure(s):** RT-PCR confirmed SARS-CoV-2 or rhinovirus infection.

**Results:** Close contact with a SARS-CoV-2 case (adjusted odds ratio, aOR 4.3, 95% CI 3.7-5.0) and loss of smell/taste (aOR 3.7, 95% CI 3.0-4.5) were the variables most associated with SARS-CoV-2 test positivity, but both attenuated during the Omicron period. Contact with a vaccinated case (aOR 2.4, 95% CI 1.7-3.3) was associated with a lower odds of test positivity than contact with an unvaccinated case (aOR 4.4, 95% CI 2.7-7.3). Sore throat was associated with Omicron infection (aOR 2.3, 95% CI 1.6-3.2) but not Delta. Vaccine effectiveness for participants fully vaccinated with a booster dose was 43% (95% CI 11-63%) for Omicron and 92% (95% CI 61-100%) for Delta.

Variables associated with rhinovirus test positivity included age <12 years (aOR 4.0, 95% CI 3.5-4.6) and reporting a runny or stuffy nose (aOR 4.6, 95% CI 4.1-5.2). Race, region, and household crowding were significantly associated with both SARS-CoV-2 and rhinovirus test positivity.

**Conclusions and Relevance:** Estimated risk factors and symptoms associated with SARS-CoV-2 infection have changed over time. There was a shift in reported symptoms between the Delta and Omicron variants as well as reductions in the protection provided by vaccines. Racial and socioeconomic disparities persisted in the third year of SARS-CoV-2 circulation and were also present in rhinovirus infection, although the causal pathways remain unclear. Trends in testing behavior and availability may influence these results.

**Key Points:** 

**Question:** What are the characteristics associated with SARS-CoV-2 and rhinovirus infection?

**Findings:** In this test-negative design study of 23,278 participants, reporting close contact with a SARS-CoV-2 case was the strongest risk factor associated with test positivity. Loss of smell and taste was associated with the Delta variant, but not the Omicron variant. Vaccination and prior infection provided greater protection against Delta infection than Omicron Infection. Young age was the strongest predictor of rhinovirus positivity. Sociodemographic disparities were present for both SARS-CoV-2 and rhinovirus.

**Meaning:** Monitoring factors associated with respiratory pathogen test positivity remains important to identify at-risk populations in the post-SARS-CoV-2 pandemic period.

## BACKGROUND

Studies from the United Kingdom demonstrate that risk factors and symptoms associated with SARS-CoV-2 infection have fluctuated over the course of the pandemic and should be reassessed periodically to guide control strategies.^1–3^ In the US, early studies identified contact with a case and community and workplace exposures as important risk factors, while many studies also noted the disproportionate impact on Black, Hispanic, and socioeconomically disadvantaged communities.^4,5,14–16,6–13^ Few studies in the US have reexamined these risk factors in the context of widespread vaccination and circulation of new variants.

A feature of the SARS-CoV-2 pandemic has been substantial reduction in the circulation of endemic respiratory pathogens because of non-pharmaceutical interventions (NPIs), with rhinovirus as a notable exception.^17–20^ After an initial decline in activity during the Spring 2020 lockdown period, rhinovirus quickly rebounded to circulate at pre-pandemic levels.^21–23^ Rhinovirus is a common pathogen that typically causes symptomatic upper respiratory tract disease in children and adults.^24^ Though it is frequently detected among hospitalized pneumonia patients,^25,26^ it is generally thought to have a mild disease course. As a result, rhinovirus is relatively poorly studied and risk factors for infection remain unclear.

Here we examine changes in risk factors and symptoms associated with SARS-CoV-2 test positivity among participants of the Greater Seattle Coronavirus Assessment Network (SCAN) Study over a 22-month pandemic period encompassing the circulation of new variants and rising immunity from natural infection and vaccination. We examine characteristics associated with all SARS-CoV-2 infections and compare risk factors for Delta and Omicron infections separately. We contrast our findings with characteristics associated with rhinovirus test positivity.

## METHODS

### Study Design

SCAN was designed as a cross-sectional surveillance study using online community recruitment to monitor the incidence of SARS-CoV-2 and other respiratory pathogens in the greater Seattle area from June 10, 2020, to April 30, 2022. Participants were largely symptomatic (>90%) and oversampled in underserved areas (see eMethods for recruitment criteria).

Enrolled participants received a free testing kit delivered to their home for self-collection of a nasal swab.^27^ Samples were tested by polymerase chain reaction (PCR) for the presence of SARS-CoV-2 and rhinovirus. Presumed Delta and Omicron variants were identified using S-gene Target Failure criteria. See eMethods for study design, exclusion criteria (eFigure 1), and laboratory methods.

### Variable definitions

To assess risk factors for infection with SARS-CoV-2 and rhinovirus, we used a test-negative design.^28–30^ Care-seeking participants who tested positive are referred to as “cases” while those testing negative serve as the control group. The dependent variables in regression models were either SARS-CoV-2 or rhinovirus positivity. Not all samples were tested for non-SARS-CoV-2 pathogens. Therefore, in the main analysis we defined a SARS-CoV-2 case as any participant with a positive SARS-CoV-2 result (including coinfections) and a SARS-CoV-2 control as any participant with a negative SARS-CoV-2 result. In sensitivity analysis we restricted the sample to those tested for other pathogens and excluded coinfections (eMethods). A similar logic was applied to presumed Delta and Omicron cases, with controls drawn from a comparable period as cases (June 1 -December 12, 2021, for Delta; December 12, 2021, onwards for Omicron). Rhinovirus cases were defined as anyone with a positive rhinovirus result (and a negative enterovirus result), excluding coinfections with SARS-CoV-2. Rhinovirus controls included participants who had a negative rhinovirus result, after excluding those positive for SARS-CoV-2.

Independent variables missing >5% of data were excluded and for the remaining variables complete cases were used. Five core sociodemographic variables were included in all models: age, sex, race/ethnicity, county region (based on Public User Microdata Area (PUMA) of residence), and social and economic risk index ([SERI], a local risk indicator based on census-tract of residence). SERI was developed by Public Health Seattle and King County (PHSKC) to describe socioeconomic inequalities and identify communities at increased risk for COVID-19 (see eMethods and^31^ for details). We included ten additional demographic and health behavior variables (see eMethods for full list), and indicator variables for 16 symptoms and number of symptoms (≤3 or >3). We used a categorical variable for time-period: Early (June 10, 2020-January 31,2021), Intermediate (February 1, 2021 – December 11, 2021), Omicron (December 12, 2021-April 30,2021). For the SARS-CoV-2 model we included the log of the weekly reported SARS-CoV-2 cases in the county as an external measure of community incidence (Figure 1a). For the rhinovirus model we included the weekly percent positive of rhinovirus tests from the Pacific Northwest Respiratory Virus Epidemiology Data.^32^

**Figure 1.**
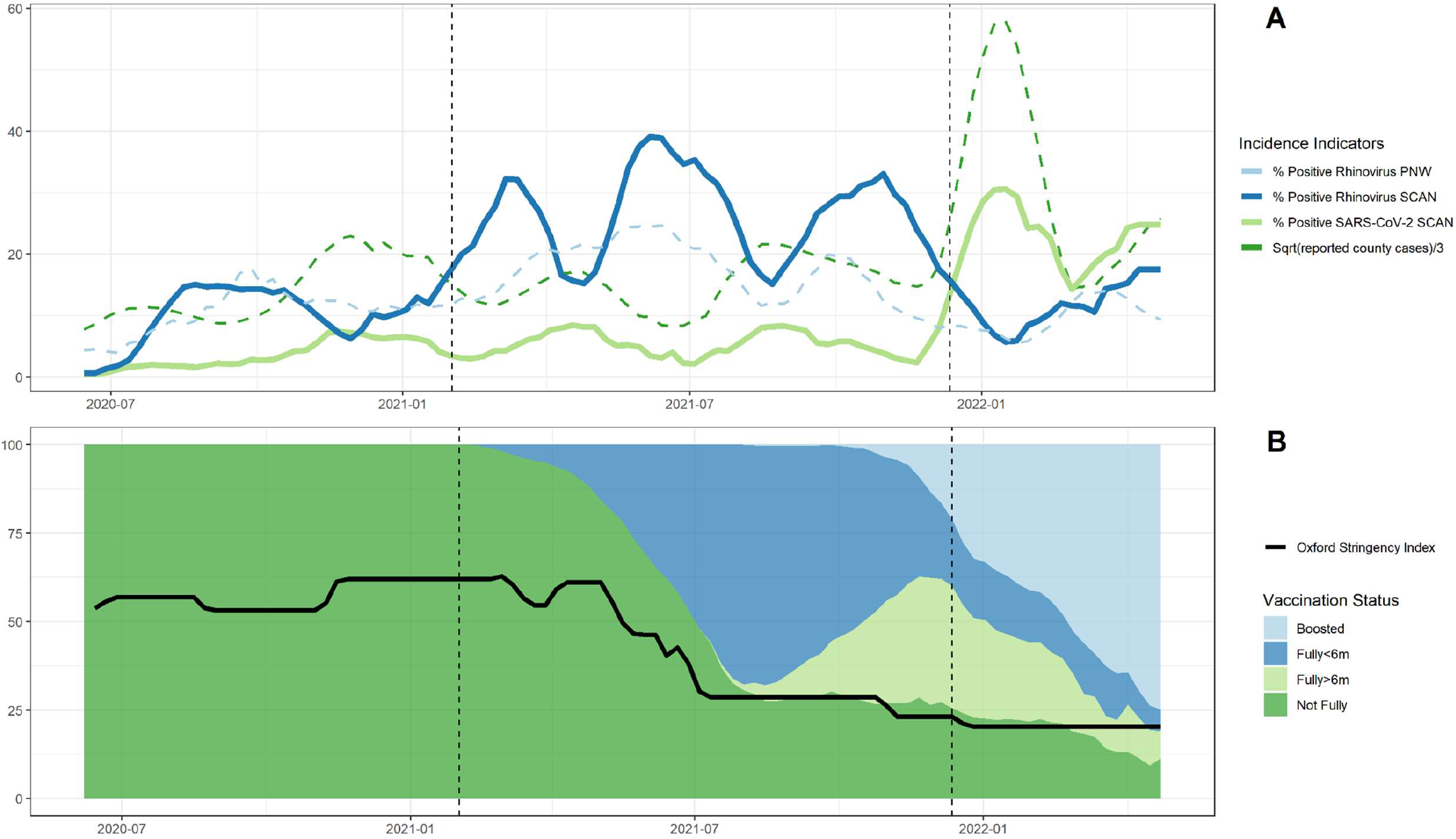
Trends in viral circulation, non-pharmaceutical interventions, and vaccination. Panel A) The 5-week moving average of the percent of samples positive for SARS-CoV-2 and rhinovirus in the SCAN study population over time, the square root of the reported SARS-CoV-2 cases in King County divided by three (for easier visualization) and the % positive of rhinovirus tests in the Pacific Northwest (PNW) surveillance system. Vertical dashed lines separate time-periods (Early, Intermediate, Omicron). Panel B) The percent of SCAN study participants by vaccination status (not fully vaccinated = unvaccinated or received an incomplete primary schedule, fully vaccinated more than 6m ago, fully vaccinated less than 6m ago, boosted (1 or more boosters)) and the weekly average of the Oxford Stringency Index in Washington State, which reflects the strength of interventions on a scale of 0 to 100%. Dashed lines separate time-periods (Early, Intermediate, Omicron).

### Statistical Analysis

Following prior work,^1,7,8,13,33^ we used univariate and multivariable logistic regression to assess associations between SARS-CoV-2 test positivity and the independent variables, and used forward and backward stepwise Akaike Information Criterion (AIC) for variable selection. To explore time trends, we tested interactions between each variable and time-period (eMethods). Confidence intervals for the adjusted model with interactions were based on non-parametric bootstrap (1000 simulations). We used the same approach with the rhinovirus model, but with SARS-CoV-2 specific variables excluded, i.e., contact with a SARS-CoV-2 case, prior SARS-CoV-2 infections, and COVID-19 vaccination. Sensitivity analyses based on choice of model structure and controls are provided in the supplement (eMethods; eTable 2; eFigure 2; eFigure 3).

### Ethics and Funding

This study was approved by the University of Washington Institutional Review Board. All participants provided informed consent at enrollment. Funding for the study was provided by Gates Ventures.

## RESULTS

### Study Population

Analyses included data from 23,278 individuals, from whom there were 1212 (5.2%) SARS-CoV-2 positive cases, including 36 coinfections with rhinovirus, and 2574 (11.1%) rhinovirus positive cases (Table 1). The median age (IQR) of participants was 34.3 years (22.4-45.1), 59.1% of participants were female, 37.7% identified as non-White, 31.4% resided in South King County, and 27.7% resided in a high-risk census tract based on SERI. Relationships between demographic variables are provided in the supplement (eFigure 2). During the Omicron period 44.8% of participants had received a booster vaccine dose (Figure 1b). Among participants reporting close contact with a case (<6 feet away for ≥10 minutes), social contacts were the most common (51.3%), followed by household (29.6%) and workplace contacts (22.7%).

**Table 1.** Demographic characteristics of SCAN participants, June 10, 2020 – April 30, 2022

|  | **Early** | **Intermediate** | **Omicron** | **Overall** |
| --- | --- | --- | --- | --- |
|  | **6/10/20 -**<br>**1/30/21** | **2/1/21 -**<br>**12/11/21** | **12/12/21 -**<br>**4/30/22** | **6/10/20 -**<br>**4/30/22** |
| **Characteristics** | **N=17158** | **N = 4733** | **N = 1387** | **N = 23278** |
| **Test Results** |  |  |  |  |
| SARS-CoV-2 cases | 579 (3.4%) | 264 (5.6%) | 369 (26.6%) | 1212 (5.2%) |
| Presumed Delta | -- | 126 (2.7%) | 8 (0.6%) | 134 (0.6%) |
| Presumed Omicron | -- | -- | 330 (22.3%) | 330 (1.4%) |
| Rhinovirus cases | 1314 (7.7%) | 1146 (24.2%) | 114 (8.2%) | 2574 (11.1%) |
| Other pathogen positive | 688 (4.0%) | 451 (9.5%) | 126 (9.1%) | 1265 (5.4%) |
| Pan-negative | 11471 (66.9%) | 2951 (62.3%) | 740 (53.4%) | 15162 (65.1%) |
| Not tested for other pathogens | 3599 (21.0%) | 129 (2.7%) | 116 (8.4%) | 3844 (16.5%) |
| **Age** |  |  |  |  |
| Median [IQR] | 35.1 [23.7,46.0] | 31.9 [15.3,42.1] | 34.8 [21.3,44.8] | 34.3 [22.4,45.1] |
| 12 – 50 years | 11918 (69.5%) | 3065 (64.8%) | 963 (69.4%) | 15946 (68.5%) |
| < 12 years | 1969 (11.5%) | 985 (20.8%) | 157 (11.3%) | 3111 (13.4%) |
| ≥ 50 years | 3271 (19.1%) | 683 (14.4%) | 267 (19.3%) | 4221 (18.1%) |
| **Sex** |  |  |  |  |
| Female | 10021 (58.4%) | 2853 (60.3%) | 880 (63.4%) | 13754 (59.1%) |
| Male | 7137 (41.6%) | 1880 (39.7%) | 507 (36.6%) | 9524 (40.9%) |
| **Race/Ethnicity** |  |  |  |  |
| White | 10963 (63.9%) | 2803 (59.2%) | 727 (52.4%) | 14493 (62.3%) |
| Asian | 2806 (16.4%) | 818 (17.3%) | 341 (24.6%) | 3965 (17.0%) |
| Black | 409 (2.4%) | 191 (4.0%) | 51 (3.7%) | 651 (2.8%) |
| Hispanic | 1584 (9.2%) | 442 (9.3%) | 137 (9.9%) | 2163 (9.3%) |
| Other | 1396 (8.1%) | 479 (10.1%) | 131 (9.4%) | 2006 (8.6%) |
| **County Region** |  |  |  |  |
| North | 12245 (71.4%) | 2972 (62.8%) | 763 (55.0%) | 15980 (68.6%) |
| South | 4913 (28.6%) | 1761 (37.2%) | 624 (45.0%) | 7298 (31.4%) |
| **SERI** |  |  |  |  |
| Low | 6946 (40.5%) | 1745 (36.9%) | 385 (27.8%) | 9076 (39.0%) |
| Moderate | 5799 (33.8%) | 1497 (31.6%) | 451 (32.5%) | 7747 (33.3%) |
| High | 4413 (25.7%) | 1491 (31.5%) | 551 (39.7%) | 6455 (27.7%) |
| **Vaccination Status** |  |  |  |  |
| Not Fully Vaccinated | 17158 (100%) | 2629 (55.5%) | 280 (20.2%) | 20067 (86.2%) |
| Fully Vaccinated (>6m ago) | 0 (0%) | 423 (8.9%) | 284 (20.5%) | 707 (3.0%) |
| Fully Vaccinated (<6m ago) | 0 (0%) | 1589 (33.6%) | 201 (14.5%) | 1790 (7.7%) |
| Boosted | 0 (0%) | 92 (1.9%) | 622 (44.8%) | 714 (3.1%) |
| **Household Size** |  |  |  |  |
| <5 people | 14224 (82.9%) | 3900 (82.4%) | 1160 (83.6%) | 19284 (82.8%) |
| ≥5 people | 2934 (17.1%) | 833 (17.6%) | 227 (16.4%) | 3994 (17.2%) |
| **Prior Test History** |  |  |  |  |
| Prior positive (>90 days ago) | 80 (0.5%) | 116 (2.5%) | 64 (4.6%) | 260 (1.1%) |
| No prior testing | 10416 (60.7%) | 1175 (24.8%) | 287 (20.7%) | 11878 (51.0%) |
| **Median Symptoms [IQR]** | 3.00 [1.0,5.0] | 3.00 [2.0,6.0] | 4.00 [2.0,7.0] | 3.00 [2.0,5.0] |

### Risk factors and symptoms associated with SARS-CoV-2 infection

In multivariable logistic regression, close contact with a case had the strongest association with a positive SARS-CoV-2 test result (aOR 4.3, 95% CI 3.7-5.0; Figure 2a, eTable 3), but this association dropped to 2.1 aOR (95% CI 1.1-3.8; Table 2) during the Omicron period. In sub-analysis accounting for vaccination status of the contact from October 2021-April 2022, we found that participants reporting contact with a vaccinated case were less likely to test positive than participants reporting contact with an unvaccinated case (aOR 2.4, 95% CI 1.7-3.3, versus aOR 4.4, 95% CI 2.7-7.3), however, this difference was not statistically significant (Table 3). Participants identifying as Black (aOR 2.0, 95% CI 1.5-2.7) or Hispanic (aOR 2.1, 95% CI 1.7-2.6) and participants living in high SERI census tracts (aOR 1.6, 1.2-2.0), households with ≥5 people (aOR 1.4, 95% CI 1.2-1.7), or reporting travel in the last 14 days had a significantly higher odds of testing positive than White participants or those living in low SERI census tracts, households with <5 people, and no travel history. Participants living in South King County and children <12 years had higher odds of testing positive than participants from North King County or participants 12-50 years during the Early period, but this was not statistically significant during later periods (Table 2). In sensitivity analysis using controls negative for all pathogens (pan-negative), the interaction between time-period and age lost significance, and the risk associated with young age was higher (aOR 1.7 [95%CI 1.2-2.3]; eFigure3), than when controls included participants positive for other pathogens. Participants who were fully vaccinated with a booster dose, self-reported a previous positive test >90 days ago, or reported attending or working at a school in the past 14 days were all significantly less likely to test positive than those who were not fully vaccinated, had never tested positive, or did not attend or work at a school. Among symptoms, loss of smell or taste (aOR 3.7, 95% CI 3.0-4.5), fever (aOR 2.4, 95% CI 2.0-2.8), and cough (aOR 2.4, 95% CI 2.0-2.8) were the strongest predictors of test positivity (Figure 2b).

**Table 2.**
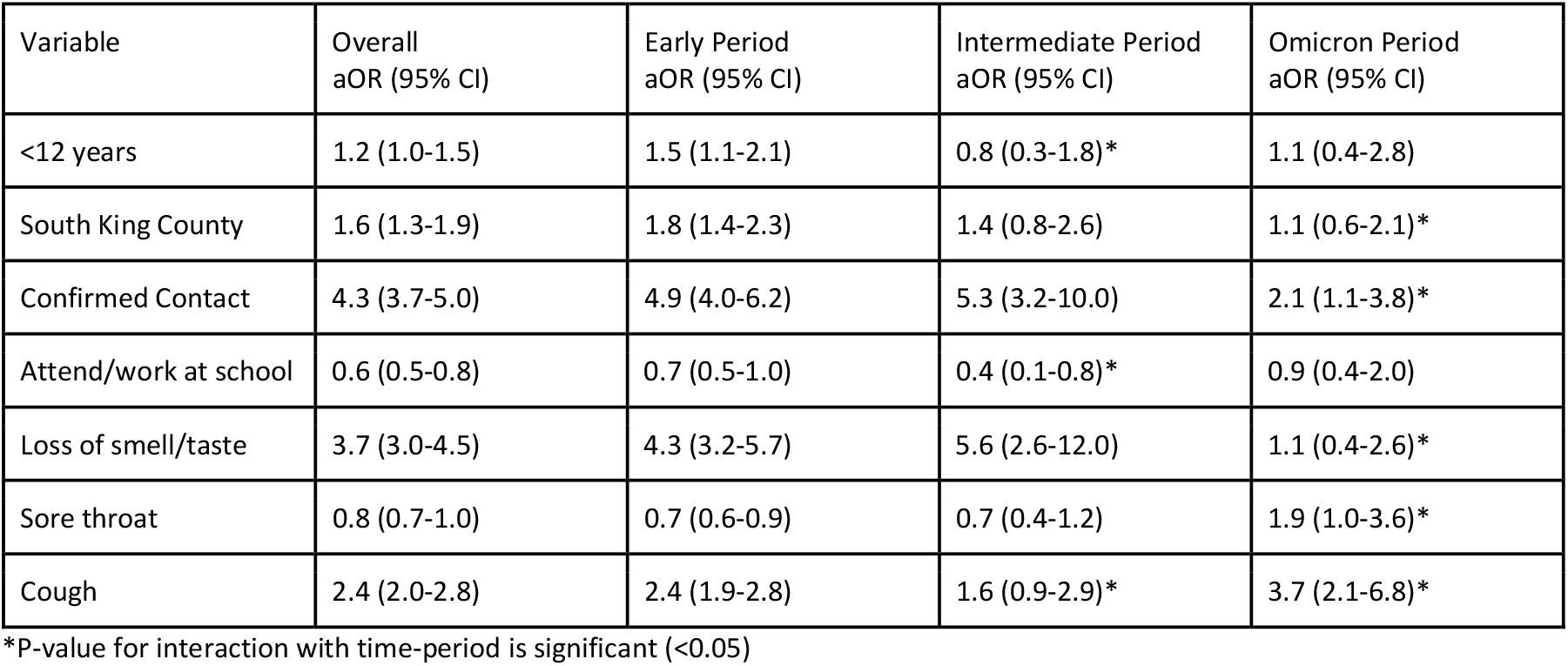
Adjusted odds ratios by time-period for variables with significant interactions

**Table 3.** Odds ratio for testing positive for participants reporting contact with a case in the past 2 weeks based on vaccination status and reported symptoms of the contact. These variables were collected for the later part of the study period, October 2021-April 2022.

| Date range | October 2021 - April 2022<sup>1,2</sup> |  | December 2021 - April 2022<sup>3,4</sup> |  |
| --- | --- | --- | --- | --- |
|  | N = 2310 |  | N = 1473 |  |
|  | OR (95% CI) | p-value | OR (95% CI) | p-value |
| No recent contact with a case (Reference) | 1.0 |  | 1.0 |  |
| Recent contact with an unvaccinated case | 4.4 (2.7-7.3) | 0.106<sup>5</sup> | 2.7 (1.6-4.8) | 0.656<sup>5</sup> |
| Recent contact with a vaccinated case | 2.4 (1.7-3.3) |  | 1.9 (1.3-2.6) |  |
| No recent contact with a case (Reference) | 1.0 |  | 1.0 |  |
| Recent contact with a symptomatic case | 3.3 (2.3-4.7) | 0.119<sup>6</sup> | 2.4 (1.7-3.5) | 0.231<sup>6</sup> |
| Recent contact with an asymptomatic case | 2.0 (1.3-2.9) |  | 1.5 (1.0-2.3) |  |

<sup>1</sup> Adjusted for age, sex, race, region, SERI, cough, sore throat, fever, loss of smell or taste, attending/working at school, prior positive test, vaccination status of participant, log(cases reported in community), dominant variant.
<sup>2</sup> Includes all cases within date range (Delta, Omicron, undetermined).
<sup>3</sup> Adjusted for age, sex, race, region, SERI, cough, sore throat, fever, loss of smell or taste, attending/working at school, prior positive test, vaccination status of participant, log(cases reported in community).
<sup>4</sup> Includes presumed Omicron cases only.
<sup>5</sup> P-value for comparison between contact not vaccinated and contact vaccinated. Calculated using Tukey's HSD.
<sup>6</sup> P-value for comparison between contact symptomatic and contact asymptomatic. Calculated using Tukey's HSD

**Figure 2.**
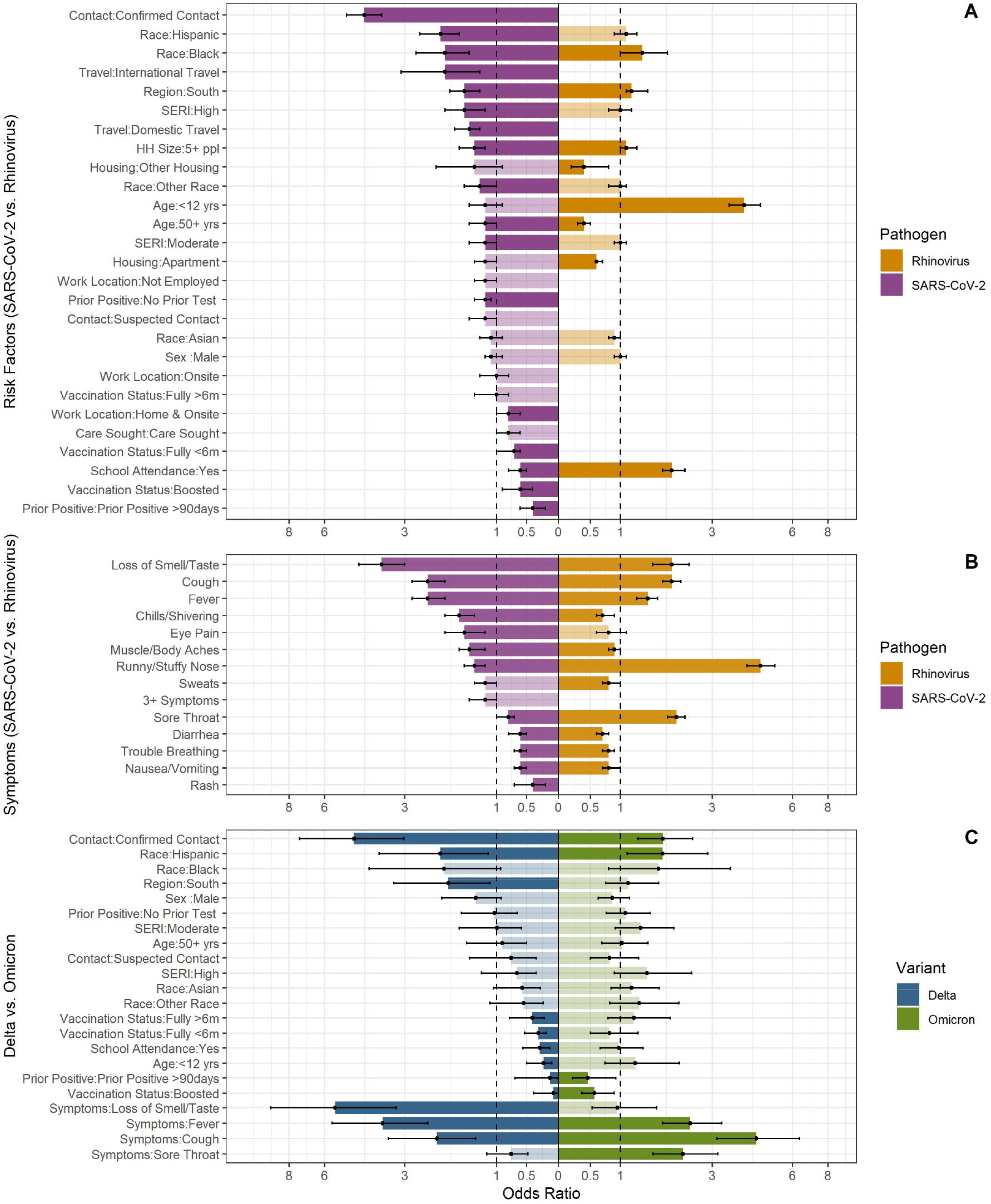
Risk factors and symptoms associated with SARS-CoV-2 and rhinovirus infection. Panel A) Adjusted odds ratios and 95% CIs for risk factors associated with SARS-Cov-2 and rhinovirus positivity. Odds ratios are ordered from highest to lowest for SARS-CoV-2. Missing bars indicate the variable was not included in the model a priori or dropped during model selection. Transparent bars and 95% CIs touching the dashed line are not statistically significant. Panel B) Same as A but for symptoms. Panel C) Same as A and B but for Delta and Omicron variants. Separate models are run for the risk of test positivity with Delta and Omicron

### Differences between Delta and Omicron variants

We found several notable differences in analysis comparing the Delta and Omicron variants (Figure 2c). Reporting contact with a case was associated with a substantially higher odds of testing positive for Delta (aOR 4.7, 95% CI 3.0-7.3) compared to Omicron (aOR 1.8, 95% CI 1.5-2.5). Similarly, loss of smell and taste was associated with an odds ratio of 5.5 (95% CI 3.3-9.3) for Delta, but was not significant for Omicron, while sore throat was associated with Omicron infection (aOR 2.3, 95% CI 1.6-3.2) but not Delta. In terms of protective factors, a self-reported previous infection was less protective against Omicron (aOR 0.5, 95% CI 0.2-0.9) than Delta (aOR 0.1, 95% CI 0.0-0.7), as was vaccination. Being fully vaccinated did not provide significant protection against Omicron but was associated with a vaccine effectiveness (VE) of 69% against symptomatic infection with Delta in the first six months (aOR= 0.31 (95% CI 0.19-0.53)), and 59% six months after vaccination (aOR=0.41 (95% CI 0.22-0.78)). An additional booster dose increased VE to 92% against Delta (aOR 0.08, 95% CI 0.00-0.39) and 43% against Omicron (aOR 0.57, 95% CI 0.37-0.89).

### Risk factors and symptoms associated with rhinovirus infection

Overall, there were fewer variables associated with rhinovirus test positivity than SARS-CoV-2 (Figure 2a, eTable 4). The greatest risk factor was age <12 years (aOR 4.0, 95% CI 3.5-4.6). Similar to SARS-CoV-2, we found that identifying as Black, living in South King County or in a household with ≥5 people were also associated with rhinovirus positivity. In contrast to SARS-CoV-2, attending or working at a school was associated with an increased odds of rhinovirus positivity (aOR 2.0, 95% CI 1.8-2.3). Reporting a runny or stuffy nose (aOR 4.6, 95% CI 4.1-5.2) or sore throat (aOR 2.1, 95% CI 1.9-2.3) (Figure 2b) were the symptoms most associated with rhinovirus test positivity. Results from sensitivity analyses and interactions with time-period are presented in the supplement (eFigure 5; eFigure 6). Interestingly, the percent positive for rhinovirus was anticorrelated with the percent positive for SARS-CoV-2 infection in our sample, with 12-14 weeks separating the peaks of each virus (eFigure 7).

## DISCUSSION

We have presented risk factors and symptoms associated with SARS-CoV-2 infection, considered how these associations have changed over a 22-month period, and compared against characteristics associated with rhinovirus test positivity. We found that the characteristics most strongly associated with SARS-CoV-2 early in the pandemic, namely contact with a case and loss of smell or taste, attenuated, or disappeared during the Omicron period, while reporting a sore throat emerged as significant. Several sociodemographic characteristics associated with SARS-CoV-2 positivity persisted over time but geographic disparities weakened. Vaccination and prior infection offered considerably more protection against the Delta variant compared to the Omicron variant and receiving a booster dose improved protection against both variants. In contrast to SARS-CoV-2, younger age and attending/working at a school were important risk factors for rhinovirus.

In line with prior studies,^4,6^ risk of SARS-CoV-2 test positivity was higher for participants reporting close contact with a case compared to those reporting no case contacts. However, the observed attenuation of this association over time, particularly during the period when Omicron predominated, is surprising given Omicron’s transmission advantage.^34–36^ It is possible that, high levels of Omicron circulation in the community and reduced NPI stringency could have resulted in many asymptomatic/pre-symptomatic or unknown contacts with cases, driving estimates towards the null. Inability to link cases has important implications for contact tracing efforts, and as of February 2022, the Centers for Disease Control and Prevention no longer recommends universal contact tracing or case investigation for COVID-19.^37^ Alternatively, high levels of vaccination during the Omicron period may have lessened the risk of onward transmission.^38–42^ We found that contact with an asymptomatic or vaccinated case lowered the odds of testing positive compared to contact with a symptomatic or unvaccinated case, though this was not statistically significant, perhaps owing to small sample size. Although vaccines are less effective against Omicron infection compared to earlier variants,^43,44^ the extent of primary vaccination and booster coverage in our study population was much higher during the period of Omicron circulation than earlier periods. Modeling studies early in the pandemic suggested that higher vaccine coverage with lower vaccine efficacy might reduce SARS-CoV-2 cases more than lower coverage with a higher efficacy vaccine.^45^ Further, there is evidence that vaccines remain effective against severe outcomes with Omicron,^46^ but SCAN data are not appropriate to study severe infections.

Our VE estimates for Delta and Omicron are consistent with other studies,^43,44,47,48^ showing waning protection against symptomatic infection with the Delta variant after 6-months and restoration of protection to >90% following an additional booster dose. We found substantially less protection from vaccination and self-reported prior infection against Omicron, consistent with the immune escape feature of Omicron. We did not adjust for time since prior infection but acknowledge this could impact results. There were also notable differences in the reported symptoms between Delta and Omicron. Loss of smell and taste, the early hallmark of SARS-CoV-2 infection, was not significantly associated with Omicron infection, while Omicron-infected individuals were more likely to report a sore throat, in line with other studies.^2,3^

Many studies have documented the disproportionate toll of COVID-19 on minority groups and economically disadvantaged communities,^7–13^ and we too found higher risk of infection in these populations. We included a combination of individual-level (i.e., race/ethnicity, occupation, and household size) and community-level (i.e., region, SERI) sociodemographic variables in our model. Limited ability to work from home or socially distance have been proposed as possible mechanisms driving racial disparities in SARS-CoV-2 infection.^49^ Onsite work was associated with SARS-CoV-2 infection in univariate analysis, but not in the adjusted model, while the associations with race/ethnicity and household crowding remained in the adjusted model and persisted in all time-periods. We were unable to include other individual-level variables that could help explain the differences observed for race/ethnicity (i.e., household income, education, comorbidities, or mask wearing and other non-pharmaceutical interventions), due to missing data. However, household income and educational attainment were factors included in the community-level SERI score (eMethods).

At the community-level, we found that geographic differences between North and South King County gradually diminished. Mixing between North and South King County increased after stay-at-home orders were lifted in June 2020, which may have homogenized risk of exposure over time. Although we did not include an interaction with time for SERI in the final model because of our stringent inclusion criterion, SERI was not a significant predictor in either the Delta or Omicron models, possibly due to small sample size. Overall, the attenuation of geographic and sociodemographic disparities in the risk of infection would be expected during SARS-CoV-2’s transition to endemicity, where early infection of high-exposure groups confers immunity to later epidemic waves, with few eventually escaping infection. Although prior infection offered less protection against Omicron than earlier variants, test-positivity was still lower among previously infected individuals. Alternatively, communities severely impacted earlier in the pandemic may be more willing to adopt behavior change to limit transmission while communities that largely escaped earlier waves may be less cautious in later periods. Of note, many sociodemographic variables are interrelated, and reconstruction of the causal pathways that affect infection and the total contribution of each variable would require different approaches than used here. Furthermore, our estimates for these sociodemographic variables are drawn from a relatively homogenous, urban study population and may not be broadly generalizable. Moreover, SERI is a localized measure, developed specifically for COVID-19, and may not apply in other settings or to other health outcomes.

Odds of SARS-CoV-2 infection in children <12 were slightly higher than for participants 12-50 years in the early time-period, but not in the later periods, despite low levels of vaccination in children even after they became eligible. We cannot rule out that changes in sampling over the study period could affect these results, however, restricting analysis to pan-negative controls removed this decreasing trend. This suggests that the rebound of rhinovirus later in the pandemic may have impacted the propensity for children to test positive for SARS-CoV-2 in our study. Accounting for virus co-circulation and potential interactions between pathogens will be an important aspect of ascertaining risk for respiratory infections in the post-pandemic period. Further, working at or attending school in the two weeks prior to enrolling in SCAN was associated with lower odds of SARS-CoV-2 test positivity, particularly during the Intermediate period which coincides with the start of the school year in Fall 2021. In general, studies have shown that NPIs and frequent testing can reduce the occurrence of outbreaks in schools.^50,51^ However, when strong interventions are in place in the work or school environment, protective associations can reflect reverse causality, a phenomenon also reported with onsite work in the UK.^1^ Children and staff allowed to attend school have to be free of symptoms and lack recent contact with a SARS-CoV-2 case, and are thus unlikely to test positive.

In contrast to SARS-CoV-2, younger age and school attendance were highly associated with rhinovirus infection. The persistent circulation of rhinovirus in children throughout the SARS-CoV-2 pandemic, despite declines in other pathogens, has been widely observed.^20–23^ Accordingly, we detected more rhinovirus than SARS-CoV-2 infections, but very few other viral pathogens. Another study found that SARS-CoV-2 may repress rhinovirus infection on individual-level data,^52^ which may explain the anticorrelation in circulation we observed at the population level. In the past, similar viral interference has been observed between rhinovirus and influenza.^53^ Few studies have considered demographic risk factors beyond age for rhinovirus infection. Rhinovirus infection was more likely in Black participants and participants living in South King County or households with ≥5 people compared to White participants and those living in North King County or households with <5 people. This was less pronounced than the differences observed for SARS-CoV-2 infection, but suggest that similar sociodemographic inequalities also modulate the burden of endemic respiratory pathogens.^54^ Examining sociodemographic risk factors for other endemic pathogens, and how they may interact with SARS-CoV-2 circulation, is an important consideration for future analyses.

### Limitations

The self-referring study participants were not a representative sample of the King County population. Eligibility criteria, testing demand, and the composition of the study participants changed over the course of the study (eMethods, Table 1). Participation in less-affluent South King County increased over time as a result of targeted efforts to broaden access to testing. This may influence the associations we identified, particularly when considering time trends. All data were based on self-report and while we expect that misclassification is minimal, we acknowledge that this may impact our results.

Misclassification could be quite important for prior infection since testing propensity was low in the early phase of the pandemic. Furthermore, prolonged shedding of rhinovirus is common, particularly when PCR testing is utilized, which may lead to an overestimate of rhinovirus infection.

## CONCLUSIONS

There have been several important changes in the risk factors and symptoms associated with SARS-CoV-2 test positivity. Reported symptoms shifted between the Delta and Omicron variants, while vaccines and previous infections offered reduced protection against later symptomatic infections. High levels of SARS-CoV-2 circulation during the Omicron period weakened the association between test positivity and known contact with a case, which has important implications for contact tracing. Importantly, racial and sociodemographic disparities identified early in the pandemic have persisted and also impact common pathogens like rhinovirus. Continued efforts to understand the drivers of respiratory virus infections in the post-pandemic period remain important to improve targeted interventions.

## Data Availability

All data produced in the present study are available upon reasonable request to the authors

## ACKNOWLEDGEMENTS SECTION

We would like to thank the SCAN study participants for their participation in this research. We would like to thank Public Health Seattle and King County for their contributions to the SCAN study and for developing the social and economic risk index. We would like to thank Tigran Avoundjian for his work developing the social and economic risk index and sharing it with us for research purposes. We would like to thank Christopher Craft, Kairsten Fay, Todd Seidelmann, Misja Ilcisin, Brenna Ehmen, Lani Regelbrugge, and Kathryn McCaffrey for their contributions to SCAN.

## Disclaimer

The findings and conclusions in this report are those of the authors and do not necessarily represent the official position of the U.S. National Institutes of Health or Department of Health and Human Services.

