## Supplemental Materials for "Trends in risk factors and symptoms associated with SARS-CoV-2 and Rhinovirus test positivity in King County, Washington: A Test-Negative Design Study of the Greater Seattle Coronavirus Assessment Network"

**Supplemental Materials**
**Table of Contents**

**eMethods**
A. Design of the Greater Seattle Coronavirus Assessment Network (SCAN) Study
B. Variable Definitions
C. Laboratory Methods
D. Exploring interactions with time-period
E. Sensitivity Analysis Using Different Regression Approaches
F. Sensitivity Analysis Using Different Control Groups
G. Sensitivity Analysis Using Weekly Incidences Instead of Time-Period for Interactions
H. SCAN Study Questionnaire

**eFigure 1.** Flowchart of inclusion and exclusion criteria and final sample size.

**eFigure 2.** Samples collected over time.

**eTable 1.** Comparison of participants who enrolled in SCAN multiple times versus participants who enrolled only once.

**eTable 2.** Comparison of the odds ratios and selected variables under different regression approaches

**eFigure 3.** Relationships between demographic variables.

**eTable 3.** Unadjusted and Adjusted Odds Ratios for SARS-CoV-2 test positivity, without interactions

**eFigure 4.** Sensitivity analysis for risk factors and symptoms associated with SARS-CoV-2 positivity using pan-negative controls and excluding asymptomatic participants.

**eTable 4.** Unadjusted and Adjusted Odds Ratios for Rhinovirus test positivity, without interactions

**eFigure 5.** Sensitivity analysis for risk factors and symptoms associated with rhinovirus positivity using pan-negative controls and excluding asymptomatic participants.

**eFigure 6.** Adjusted odds ratios for rhinovirus test positivity for each time-period for variables with significant interactions with time-period.

**eFigure 7.** Cross-correlation between rhinovirus and SARS-CoV-2 test positivity in the SCAN study.

**eReferences**

**eMethods**

Here we provide a more detailed description of our methodology including the study design, laboratory methods, variable definitions, and various sensitivity analyses.

#### A) Study Design

The Greater Seattle Coronavirus Assessment Network (SCAN) was launched in partnership with Public Health Seattle and King County (PHSKC) on March 23, 2020, to monitor circulation of SARS-CoV-2 in the community. The study was briefly paused on May 12, 2020, for Food and Drug Administration regulatory review and relaunched on June 10, 2020. The surveillance study is funded by Gates Ventures and ended on July 31, 2022; here we used data available from June 10, 2020, through the end of April 2022.

SCAN used multiple avenues to recruit participants from the King County area including social media advertising and community outreach through partner organizations. Eligibility criteria changed slightly over the course of the study in response to testing demand and were based on Public Use Microdata Area (PUMA) and reported symptoms (new or worsening fever, cough, or shortness of breath within the past seven days). Each PUMA had a daily allocation of enrollments with over sampling in the less-affluent Southern part of King County to ensure access and equity to testing across the county population. After an initial online screening questionnaire, participants deemed eligible based on residence and presence of symptoms at the time of screening were prompted to complete a detailed demographic and health behavior questionnaire. Enrolled participants received a free PCR-based testing kit delivered to their home for self-collection of a nasal swab.

Beginning in June of 2020, the study accommodated daily enrollments of up to 160 per day. Enrollments peaked at 250 per day between July 4, 2020, and November 17, 2020, at which time enrollments were gradually reduced to 100 per day with a PUMA allocation of 50:50 percent ratio between North and South King County. On January 26, 2021, enrollments were adjusted to 50 online community enrollments per day and the daily PUMA allocation was set at a 40:60 percent ratio between North and South King County.

SCAN participants were predominantly symptomatic (>90%), although symptoms quotas changed over time to accommodate testing needs. Symptomatic enrollees were defined as SCAN participants who self-reported experiencing either a new or worsening fever, cough, or shortness of breath within the past seven days. Asymptomatic enrollees were defined as individuals self-reporting none of these symptoms. A 75:25 percent ratio of symptomatic: asymptomatic participants was used until June 23, 2020. From June 24, 2020, through December 27, 2021, the ratio of symptomatic to asymptomatic enrollments was adjusted to 95:5 to accommodate the high demand for testing when testing options were limited. On December 28, 2021, the enrollment criteria eliminated the asymptomatic option completely as a direct result of the increased testing demand in response to the Omicron wave. Overall, 92.5% of participants in the Study were categorized as symptomatic enrollments.

In addition to online community enrollments, some participants were invited to participate as part of PHSKC's contact tracing efforts and collaborations with community-based organizations, however these samples were excluded from the analysis. We also excluded participants residing outside of King County and those reporting a positive SARS-CoV-2 test within the last 90 days, to avoid catching the later parts of long-lasting SARS-CoV-2 episodes, which would make it difficult to identify predictors of the early infection process. Exclusion criteria and sample size is shown in eFigure 1. To maintain independence between observations, each participant was included in analysis only once. For participants enrolling more than once, we used the first positive SARS-CoV-2 or rhinovirus test or most recent negative SARS-CoV-2 test, after excluding observations with missing data. Five participants tested positive for SARS-CoV-2 through SCAN more than once with an interval greater than 90 days, and of participants who enrolled multiple times, more than 90% tested negative for SARS-CoV-2 at every enrollment. Using the most recent negative test increased the sample size in the later periods of the study when daily quotas were lowered and enrollment declined. A table comparing participants who enrolled multiple times to participants who enrolled only once is provided in eTable 1.

#### B) Variable Definitions

Five core sociodemographic variables were included in all models, including 3 individual-level variables: age (<12 years, 12-50 years [ref], ≥50 years), sex (female [ref], male), race/ethnicity (White [ref], Asian, Black, Hispanic, Other); and 2 geographic variables addressing local risk in the community: region of the county (North [ref], South), and social and economic risk index (SERI). SERI was developed by PHSKC, using a combination of variables from the 2015-2019 American Community Survey, as a geographic indicator to describe socioeconomic inequalities and identify communities at increased risk of COVID-19.[1] With SERI, census tracts are given a score by summing the values from eight unweighted variables including: 1) the percentage of the population identifying as people of color; 2) the percentage of limited English-speaking households; 3) the percentage of the population born outside of the united states; 4) the median number of occupants per household; 5) the percentage of adults in essential healthcare-related occupations; 6) the percentage of adults in essential non-healthcare occupations; 7) the percentage of adults age 25+ with less than a college degree; and 8) the percentage of households less than 200% of the federal poverty level. These scores are standardized to a range between 0 and 1 and divided into three risk levels: low [ref], moderate, or high. For each participant, a SERI category was assigned based on the reported census tract of residence.

Additional independent variables analyzed included recent contact with a SARS-CoV-2 case (confirmed, suspected, none[ref]), vaccination status of the contact (added in October 2021: vaccinated, not vaccinated, don't know), symptom status of the contact (added October 2021: symptomatic, asymptomatic, don't know), vaccination status of the participant (Added in January 2021: not fully [ref], fully; Added in October 2021: boosted), household size (<5 persons per household [ref], ≥5 persons), housing type (house [ref], apartment, other), work location (home [ref], home and onsite, onsite, not employed), recent history of international or domestic travel (yes, no[ref]), care seeking for current illness (yes, no[ref]), self-reported prior test history (no prior positive test [ref], prior positive <90 days ago (excluded), prior positive >90 days ago, never tested). We also collected careful data on symptoms experienced in the last 7 days based on 16 indicator variables (loss of smell or taste, fever, cough, chills/shivering, headaches, ear pain/discharge, runny/stuffy nose, sore throat, muscle/body aches, fatigue, sweats, diarrhea, increased trouble breathing, nausea/vomiting, rash, and eye pain), and an indicator for experiencing more than three symptoms.

We categorized participants based on the number of vaccine doses and time since last vaccination. Participants were considered fully vaccinated 14 days after their second dose of an mRNA vaccine or a single dose of the Johnson & Johnson vaccine. Individuals were considered boosted 14 days after they received an additional mRNA vaccine following their primary vaccine series. Few participants remained unvaccinated in King County in the period where we were interested in vaccine effects, particularly during the Delta and Omicron waves. Therefore, participants who did not complete their primary schedule, or remained unvaccinated, were pooled into the "not fully vaccinated" category. We also categorized participants who completed a primary series without a boosted dose based on whether their last vaccination dose was less or more than 6 months prior to study enrollment.

We used a categorical variable for time-period: Early (June 10, 2020-January 31,2021), Intermediate (February 1, 2021 – December 11, 2021), Omicron (December 12, 2021-April 30,2021). The Early period represents the period before variant circulation and vaccination. Vaccination started for adults ≥65 years in King County in December 2020 but was not common among our study participants until after January 2021 (Figure 1). The intermediate period represents the circulation of multiple variants (with Delta becoming dominant in summer/fall 2021) and the ramp up of vaccination. The Omicron period is characterized by widespread vaccination and dominance of the Omicron variant. We chose not to divide the Intermediate period into multiple variant-specific periods out of concern for sample size and because several variants co-circulated in King County during spring and summer 2021.[2] We obtain Delta specific estimates through a separate regression model.

##### C) Laboratory Methods

Laboratory methods have been described elsewhere.[3,4] Participants received self-swabbing kits at home within 24 hours of requesting a test via a special carrier.[5] After swabbing, participants shipped their self-collected swabs to the Brotman Baty Institute for Precision Medicine, Seattle, Washington for laboratory testing within 48-72/hours of swabbing and 10/days of symptoms onset. Samples were tested on a Clinical Laboratory Improvement Amendments-compliant laboratory designed PCR test for the presence of SARS-CoV-2 on 2 multiplex assays (total of 4 RT-PCR reactions) targeting the Orf1b and S genes. A positive SARS-CoV-2 result had Ct values <40 on 3 of the 4 targets.

We assigned the putative variant in each swab based on the response to the S-gene. A sample was deemed "S-gene Target Failure" (SGTF) if the mean Ct for the Orf1b target was <30 and the difference between the mean Orf1b Ct and the mean S-gene Ct was ≥6 or the mean Ct for the Orf1b target was <30 and the Ct values for the S-gene targets were undetermined. A sample was deemed S-gene Target Return (SGTR) if the mean Ct for the Orf1b targets was <33, and the difference between the mean Orf1b Ct and the mean S-gene Ct was ≤3 (no undetermined Ct values). SGFT specimens collected during June-December 2021 were presumed Delta, while SGTR specimens collected after November 1, 2021, were deemed Omicron.

A majority of samples (83.5%) was also tested for 24 other respiratory pathogens (eFigure 2), by TaqMan RT-PCT on the OpenArray platform (Thermo Fisher). The OpenArray platform includes primers for two clades of rhinovirus (2 RT-PCR reactions per primer). The rhinovirus primers on the OpenArray platform detect both rhinovirus and enterovirus. Additionally, there is a separate primer for enterovirus alone. For a positive rhinovirus result, both Crt values for at least one rhinovirus primer set must be <28, with an Amp Score >1.0, and a CQ Confidence >0.5. Samples with a positive enterovirus result, in addition to a positive rhinovirus, were considered positive for enterovirus and were excluded from the analysis because we could not rule out the possibility of coinfections with rhinovirus.

**D) Exploring interactions with time-period**

To explore changes in risk factors over time, we tested interactions one at a time between each variable in the adjusted model (after variable selection) and time-period, with the Early time-period as the reference. We used a significance threshold of 0.002 (adjusting for multiple comparisons, 0.05/31 variables tested). Interactions meeting this criterion were included in the model together and were retained if they remained significant at p<0.05. We were not able to test the interaction between time-period and vaccination status as no participants were fully vaccinated during the reference time-period. Confidence intervals for the adjusted model with interactions were based on non-parametric bootstrap (1000 simulations).

**E) Sensitivity analyses using different regression approaches:**

In the main analysis, we used multivariable logistic regression with AIC selection to assess risk factors for SARS-CoV-2 and rhinovirus test-positivity. In sensitivity analyses, we also considered two complementary approaches for variable selection: LASSO regression using the glmnet package[6] and Bayesian Model Averaging using the BMA package.[7] The results of the three approaches are compared in eTable 2. For our LASSO regression model, we selected the optimal regularization parameter (lambda) through 10-fold cross-validation, and we report covariates retained by the model when using lambda.min (the lambda of the minimum mean cross-validated error) and lambda.1se (the largest value of lambda such that error is within 1 standard error of the minimum mean error). We find that the

LASSO regression model using lambda 1se (i.e., the most regularized model) retained the fewest variables, with coefficients for retained variables closer to 0 (1 after exponentiation) than the other models. This is expected as the LASSO model has the strongest stringency for variable inclusion; yet the variables retained by LASSO and their coefficients were consistent with those of the key risk factors identified in stepwise logistic regression. Bayesian Model Averaging also retained fewer variables than the stepwise regression, and similarly, coefficients for the retained variables were consistent between the two methods.

**F) Sensitivity Analysis Using Different Control Groups**

For both SARS-CoV-2 and rhinovirus, we conducted sensitivity analyses that excluded any cases with coinfections and restricted the controls to participants tested for 24 additional pathogens and negative for all (pan-negative). The rationale being that participants who were negative for SARS-CoV-2 or rhinovirus but positive for other pathogens may share risk factors and including them in the control group may dilute the estimated odds ratios. To further standardize our sample and account for changes in the proportion of asymptomatic participants, we ran additional sensitivity analyses that restricted the samples to participants reporting at least one symptom. Results from these sensitivity analyses were broadly similar to the main analysis and are provided in eFigure 2 and eFigure 3.

**G) Sensitivity Analysis Using Weekly Incidence Instead of Time-Period for Interactions**

To explore if the interaction between contact with a SARS-CoV-2 case and time-period was a function of increased circulation in the community, resulting in a greater likelihood of unknown contacts, we tested the interaction between contact with a case and our external measure of community incidence (the log of the weekly cases reported in the community). We found a significant interaction ($p<0.0001$) indicating that for each unit increase in community incidence the odds ratio associated with contact with a case decreased by 56%. To disentangle the correlation between community incidence and the Omicron variant, we also tested the interaction between community incidence and loss of smell and taste. This interaction was not statistically significant. This suggests that the changes we see in symptom presentation are driven by differences in variants rather than high levels of circulation.

**H) SCAN Study Questionnaire**

The Study questionnaire was updated several times throughout the study to reflect changes in behavioral recommendations and interventions such as hygiene and vaccination. The version of the questionnaire provided here is the most recent version of the survey, reflecting changes made on October 5, 2021. We only included variables in the model if they were consistently asked throughout the study period and were missing <5% of data.

### Illness Questionnaire

Record ID ______________________________

Hello [participant_first_name]!

Please complete this three-part survey.

Part 1: Questionnaire Part 2: Activate your SCAN Kit Part 3: Nasal Swab Collection   A link to Part 2 and Part 3 of the survey will be emailed to you once we ship your SCAN Kit. You should complete Part 2 and Part 3 when you receive your SCAN Kit in the mail.

Part 1: Questionnaire

| | |
|---|---|
| Why are you participating in SCAN/ having a COVID-19 test today? | ☐ I believe I have been exposed<br>☐ I have symptoms<br>☐ I plan to travel/be near high-risk individuals/attend an event in the next week<br>☐ I have other reasons to get tested |
| Who are you completing this survey for? | ☐ Myself<br>☐ A member of my household |
| Have you been sick in the past week with a new fever, new or worsening cough, or new or worsening shortness of breath? | ◯ Yes<br>◯ No |
| Which new or worsening symptoms have you experienced in the last 7 days? Pick any that answer the question. | ☐ I don't have any symptoms<br>☐ Feeling feverish<br>☐ Headache<br>☐ Cough<br>☐ Chills or shivering<br>☐ Sweats<br>☐ Sore throat or itchy/scratchy throat<br>☐ Nausea or vomiting<br>☐ Runny or stuffy nose<br>☐ Feeling more tired than usual<br>☐ Muscle or body aches<br>☐ Increased trouble with breathing<br>☐ Diarrhea<br>☐ Rash<br>☐ Ear pain or ear discharge<br>☐ Eye pain<br>☐ Loss of smell or taste |
| When did you first notice your current illness? Select the date on the calendar.<br><br>Note: Today's date is highlighted in yellow on the calendar. | ______________________________<br>(MM-DD-YYYY) |
| Today | ______________________________ |

Future dates are not allowed. Please change the date on the calendar to a past date to continue with the survey.

| | |
|---|---|
| Have you sought clinical care for your current illness since it started? Select all that apply. | ☐ I haven't sought care or am not currently sick<br>☐ Yes, in person at a Doctor's Office or Urgent Care<br>☐ Yes, in person at a Pharmacy (Drugstore)<br>☐ Yes, in person at a Hospital or Emergency Department<br>☐ Yes, via phone or internet<br>☐ Yes, Other |
| Have you ever been tested for COVID-19? | ◯ Yes<br>◯ No |
| Have you ever tested positive for COVID-19? | ☐ Yes, I tested positive with a nose, mouth, or throat swab<br>☐ Yes, I tested positive for antibodies with a blood sample<br>☐ No<br>☐ I don't know<br>(Select all that apply.) |
| What was the date of your most recent positive COVID-19 swab test? | ________________________<br>(If you cannot remember the exact date, please estimate.) |
| What type of COVID-19 test(s) have you had? | ☐ Nose swab through the SCAN Study or Seattle Flu Study<br>☐ Nose, mouth, or throat swab from a clinic, hospital, workplace, or other<br>☐ Antibody test from a blood sample<br>☐ Other<br>(Select all that apply.) |
| About how many times have you been tested for COVID-19? | ________________________ |
| What was the result of your most recent test? | ◯ Positive<br>◯ Inconclusive<br>◯ Negative<br>◯ I don't know |
| In the past 2 weeks, have you been in close contact with someone who tested positive for COVID-19? Close contact means that you were less than 6 feet away for at least 10 minutes. | ☐ No, I have not been in close contact with anyone that tested positive for COVID-19<br>☐ Yes, a member of my household tested positive<br>☐ Yes, I have been in close contact with a co-worker that tested positive<br>☐ Yes, I have been in close contact with a friend or person I know that tested positive<br>☐ Maybe, I was in close contact with someone with COVID-19 symptoms, but I don't know if that person has been tested<br>☐ Maybe, I was in close contact with someone with COVID-19 symptoms and was tested, but I don't yet know the test result<br>☐ I don't know<br>☐ Prefer not to say<br>(Select all that apply.) |

| | |
|---|---|
| If you have been in contact with someone who tested positive for COVID-19 in the past 2 weeks, was that person symptomatic at the time? | ◯ Yes<br>◯ No<br>◯ I don't know<br>◯ Prefer not to say |
| If you have been in contact with someone who tested positive for COVID-19 in the past 2 weeks, was that person vaccinated for COVID-19? | ◯ Yes<br>◯ No<br>◯ I don't know<br>◯ Prefer not to say |
| Have you been in contact with anyone experiencing respiratory symptoms who tested negative for COVID-19? | ◯ Yes<br>◯ No<br>◯ I don't know<br>◯ Prefer not to say |
| In the past 14 days, have you attended any community event or social gathering, such as a party, wedding, funeral, sporting event, festival, rally, or performance venue? | ◯ Yes, with 1-2 others who do not live with me<br>◯ Yes, with 3-5 others who do not live with me<br>◯ Yes, with 6-10 others who do not live with me<br>◯ Yes, with 11-20 others who do not live with me<br>◯ Yes, with more than 20 others who do not live with me<br>◯ No<br>◯ I don't know<br>◯ Prefer not to say |
| Think back to the last time you socialized with friends or family you don't live with. Which precautions did you take? Select all that apply. | ☐ I wore a facemask<br>☐ I stayed outdoors<br>☐ I ventilated indoor spaces (opened windows/doors, used fans)<br>☐ I did not share food or drinks with others<br>☐ I quarantined for 14 days before the social event<br>☐ I got tested for COVID-19 1-3 days before the social event<br>☐ I stayed 6 feet apart from others<br>☐ I washed my hands with soap or used hand sanitizer before, during, and after the social event<br>☐ I cleaned and disinfected frequently touched items and surfaces<br>☐ I only socialize with individuals who are fully vaccinated<br>☐ None of the above<br>☐ Prefer not to say |
| Do you use any of the following products (either indoors or outdoors)? | ☐ Tobacco products (e.g. cigarettes, cigars, pipes)<br>☐ Electronic cigarettes/vapor pens<br>☐ Prefer not to say<br>☐ None of the above<br>(Select all that apply.) |

| | |
|---|---|
| Do you have any of the following underlying medical conditions? Select all that apply | ☐ Cancer<br>☐ Chronic kidney disease<br>☐ COPD (chronic obstructive pulmonary disease)<br>☐ Down Syndrome<br>☐ Heart conditions, such as heart failure, coronary artery disease, or cardiomyopathies<br>☐ Immunocompromised state from solid organ transplant<br>☐ Immunocompromised state from blood or bone marrow transplant, immune deficiencies, HIV, use of corticosteroids, or other immune-weakening medications<br>☐ overweight or obese (body mass index [BMI] of 25kg/m^2 or higher)<br>☐ Sickle Cell Disease<br>☐ Type 1 diabetes mellitus<br>☐ Type 2 diabetes mellitus<br>☐ Moderate-to-severe asthma<br>☐ Cerebrovascular disease (affects blood vessels and blood supply to the brain)<br>☐ Cystic fibrosis<br>☐ Hypertension or high blood pressure<br>☐ Neurologic conditions, such as dementia<br>☐ Liver disease<br>☐ Pulmonary fibrosis (having damaged or scarred lung tissues)<br>☐ Thalassemia (a type of blood disorder)<br>☐ None of the above<br>☐ Prefer not to say<br>☐ I don't know |
| Have you received this season's influenza (flu) vaccine (since July 1, 2021) ? This includes both flu mist nasal spray and the flu shot. | ◯ Yes<br>◯ No<br>◯ Do not know |
| If completing for a child < 8 years old, did the child receive 2 doses of the influenza vaccine? | ◯ Yes<br>◯ No<br>◯ I don't know<br>◯ Prefer not to say<br>◯ Not applicable |
| If you are 65 or older, did you receive the high-dose influenza vaccine? | ◯ Yes<br>◯ No<br>◯ I don't know<br>◯ Prefer not to say<br>◯ Not applicable |
| How did you receive the flu vaccine this season (since July 1, 2021)? | ◯ Injection (flu shot)<br>◯ Nasal spray (flu mist) |

| | |
|---|---|
| What month did you get the flu shot or flu mist nasal spray this season (since July 1, 2021)? | ◯ July 2021<br>◯ August 2021<br>◯ September 2021<br>◯ October 2021<br>◯ November 2021<br>◯ December 2021<br>◯ January 2022<br>◯ February 2022<br>◯ March 2022<br>◯ April 2022<br>◯ May 2022<br>◯ June 2022<br>◯ Do not know |
| Did you receive the flu vaccine last season? (July 1, 2020 - July 1, 2021) | ◯ Yes<br>◯ No<br>◯ I don't know<br>◯ Prefer not to say |
| Has anyone in your household not received this year's influenza vaccine (since July 1, 2021)? | ◯ Yes<br>◯ No<br>◯ I don't know<br>◯ Prefer not to say |
| Have you received a COVID-19 vaccine since December 1, 2020? | ◯ Yes<br>◯ No<br>◯ I don't know<br>◯ Prefer not to say |
| How many doses (shots) of the COVID-19 vaccine did you receive? | ◯ 1 shot<br>◯ 2 shots<br>◯ 3 shots<br>◯ I do not know<br>◯ Prefer not to say |
| What is the manufacturer's name of the first dose of the COVID-19 vaccine that you received? | ◯ Pfizer<br>◯ Moderna<br>◯ Novavax<br>◯ Johnson & Johnson / Janssen<br>◯ AstraZeneca<br>◯ Other<br>◯ I don't know<br>◯ Prefer not to say |
| Please specify: | ______________________ |
| When did you receive your first dose of the COVID-19 vaccine? If you do not know the exact date, please estimate. | ______________________ |
| What is the manufacturer's name of the second dose of the COVID-19 vaccine that you received? | ◯ Pfizer<br>◯ Moderna<br>◯ Novavax<br>◯ Johnson & Johnson / Janssen<br>◯ AstraZeneca<br>◯ Other<br>◯ I don't know<br>◯ Prefer not to say |

| | |
|---|---|
| Please specify: | ________________ |
| When did you receive your second dose of the COVID-19 vaccine? If you do not know the exact date, please estimate. | ________________ |
| What is the manufacturer's name of the third dose of the COVID-19 vaccine that you received? | ◯ Pfizer<br>◯ Moderna<br>◯ Novavax<br>◯ Johnson & Johnson / Janssen<br>◯ AstraZeneca<br>◯ Other<br>◯ I don't know<br>◯ Prefer not to say |
| Please specify: | ________________ |
| When did you receive your third dose of the COVID-19 vaccine? If you do not know the exact date, please estimate. | ________________ |
| Is anyone in your household not vaccinated for COVID-19? | ◯ Yes<br>◯ No<br>◯ I don't know<br>◯ Prefer not to say |
| In the past 2 weeks, have you been able to work from home? | ◯ I am not currently employed<br>◯ No, I always have to be on-site for work<br>◯ Yes, I have only worked from home<br>◯ I have worked both from home AND on-site<br>◯ Prefer not to say |
| What industry best describes your work? | ☐ Grocery store<br>☐ Pharmacy<br>☐ Restaurant/bar<br>☐ Non-grocery retail store<br>☐ Warehouse/shipping center<br>☐ Government/public service<br>☐ Delivery driver (food)<br>☐ Delivery driver (parcel or package)<br>☐ Public transportation/airline/airport<br>☐ School or childcare<br>☐ Long-term care facility<br>☐ Healthcare setting<br>☐ Assisted living facility<br>☐ Other<br>☐ Prefer not to say<br>(Select all that apply.) |
| What other industry best describes your work? | ________________ |

| | |
|---|---|
| In the last 14 days, have you worked/volunteered in, or otherwise visited any of the following locations? Select all choices that are true for you. | ☐ Restaurant/cafe<br>☐ Bar/music venue<br>☐ Gym/exercise facility<br>☐ Salon/barber shop<br>☐ Grocery store<br>☐ Other (non-grocery) shopping establishment<br>☐ School<br>☐ Childcare Facility<br>☐ Adult long-term care facility<br>☐ Doctor's office, or healthcare facility (excluding long-term care)<br>☐ Essential workplace (excluding healthcare)<br>☐ Other workplace<br>☐ Place of worship<br>☐ Public transportation (bus, light rail, train)<br>☐ Airport<br>☐ None of the above |

**Recent Travel**

| | |
|---|---|
| In the past 14 days, have you visited a country other than the US? | ○ No, I have not traveled outside the US<br>○ Yes |
| In the past 14 days have you traveled to a US state other than Washington State? | ○ No, I have not traveled outside Washington State<br>○ Yes |

| Demographic Information | |
|---|---|
| What was your sex at birth? | ◯ Male<br>◯ Female<br>◯ Other (please specify)<br>◯ Prefer not to say |
| What is your current gender identity? | ◯ Cisgender man<br>◯ Cisgender woman<br>◯ Genderqueer/non-binary<br>◯ Man<br>◯ Transgender man/trans man<br>◯ Transgender woman/trans woman<br>◯ Woman<br>◯ Another gender not listed (please specify)<br>◯ Prefer not to answer |
| What is your current gender identity? Please specify. | __________________ |
| What is your sex? Please specify. | __________________ |
| Are you currently pregnant? | ◯ Yes<br>◯ No<br>◯ Prefer not to say |
| Are you Hispanic or Latina/Latino/Latinx? | ◯ Yes<br>◯ No<br>◯ Prefer not to say |
| How would you describe your race? | ☐ American Indian or Alaska Native<br>☐ Asian<br>☐ Native Hawaiian or other Pacific Islander<br>☐ Black or African American<br>☐ White<br>☐ Other<br>☐ Prefer not to say<br>(Select all that apply.) |
| Please choose the range that best represents your household income last year (before taxes). If you are still considered a dependent for tax purposes, choose the range that describes your parent/legal guardian's household income. | ◯ Less than or equal to $25,000<br>◯ Between $25,001 to $50,000<br>◯ Between $50,001 to $75,000<br>◯ Between $75,001 to $100,000<br>◯ Between $100,001 to $125,000<br>◯ Between $125,001 to $150,000<br>◯ Over $150,000<br>◯ Don't know<br>◯ Prefer not to say |

| | |
|---|---|
| What is your highest level of education? | ◯ Child under 5 years old<br>◯ Currently in elementary school<br>◯ Currently in middle school<br>◯ Some high school or currently in high school<br>◯ Graduated high school/obtained GED<br>◯ Some college (including vocational training, associate's degree)<br>◯ Bachelor's degree<br>◯ Advanced degree (Master's ,doctorate, medical degree, or other post-graduate educational degree)<br>◯ Prefer not to say |
| Where do you live? | ◯ House/condo/townhouse<br>◯ Homeless Shelter<br>◯ Apartment<br>◯ Dormitory<br>◯ Assisted or senior living facility<br>◯ Skilled nursing center<br>◯ Long term care or rehab facility<br>◯ Adult family home<br>◯ Inpatient or behavioral health residential center<br>◯ Permanent supportive or transitional housing center<br>◯ Correctional facility<br>◯ I don't stay in the same place every night<br>◯ Other |
| Including yourself, how many people share your kitchen or living space? | ◯ I live by myself<br>◯ 2 people<br>◯ 3 people<br>◯ 4 people<br>◯ 5 people<br>◯ 6 or more people |
| Do you have anyone in your household < 5 years old? | ◯ Yes<br>◯ No<br>◯ Prefer not to say |
| Do you have anyone in your household 5 to 12 years old? | ◯ Yes<br>◯ No<br>◯ Prefer not to say |
| Is there a reason you chose at-home testing through SCAN over other testing services or locations? Select all that apply. | ☐ I prefer to get tested from home<br>☐ It's free<br>☐ It's difficult to get an in-person appointment for testing<br>☐ I feel like I'm contributing to scientific research<br>☐ Other: _____<br>☐ I prefer not to say |
| Illness Questionnaire Date | ____________________ |
| Illness Questionnaire Date and TIME | ____________________ |

REMINDER: We are going to send a link to activate your SCAN Kit by [text_or_email] when you receive it. Please be on the lookout for this [text_or_email], as you will need to enter your sample barcode in the survey when you complete your SCAN Kit. Your sample barcode will be on your SCAN Kit box and on your sample tube. Please make sure that you do not mail your SCAN Kit back to us until you have completed the Kit Activation survey, entered in your sample barcode, and written your first and last name on your sample tube. If you do not, we may be unable to test your sample.

**eFigure 1.** Flowchart of inclusion and exclusion criteria and final sample size.

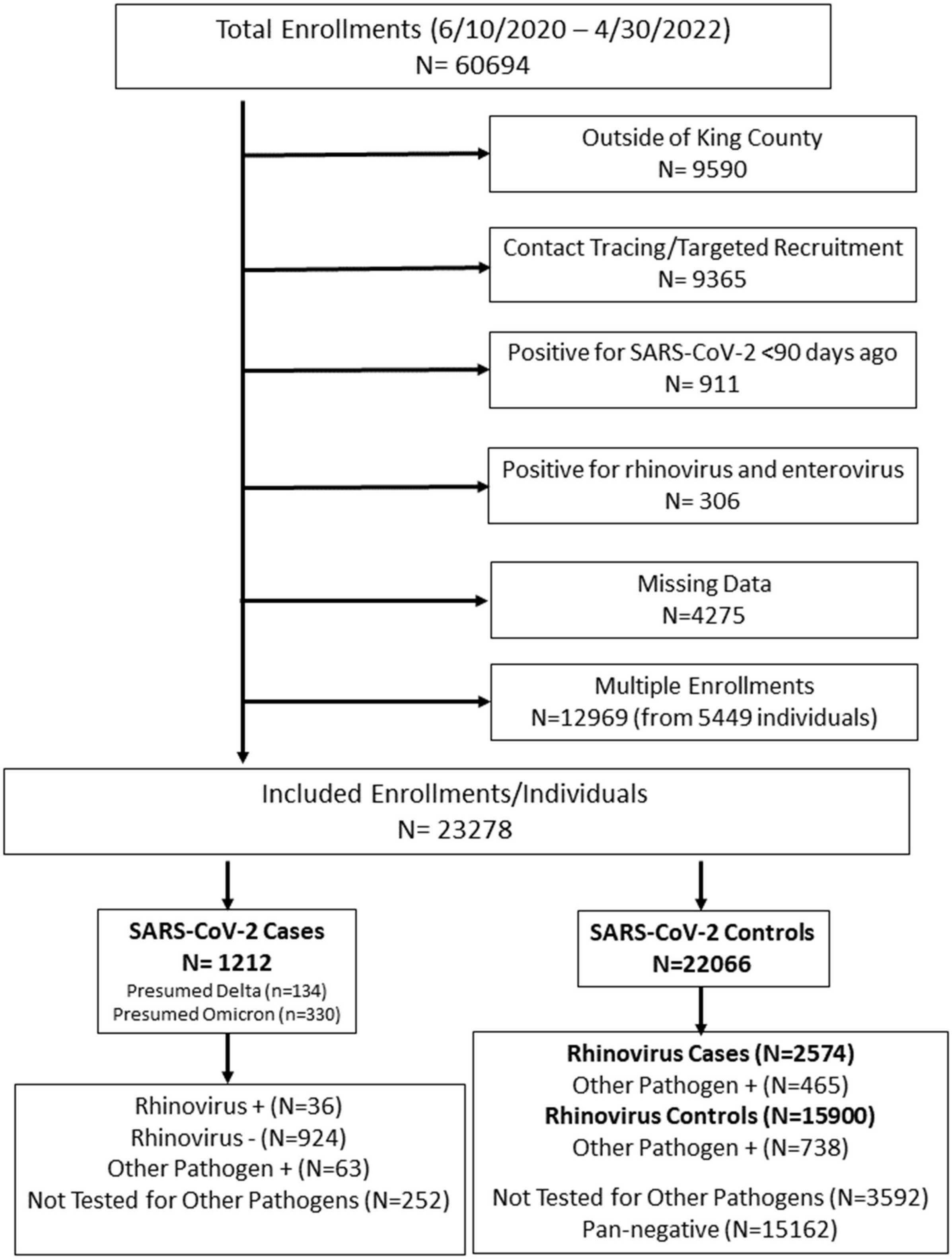

**eFigure 2.** Samples collected over time. Bars are color coded to represent the number of samples tested for SARS-CoV-2 only and those tested for SARS-CoV-2 and 24 other respiratory pathogens including rhinovirus.

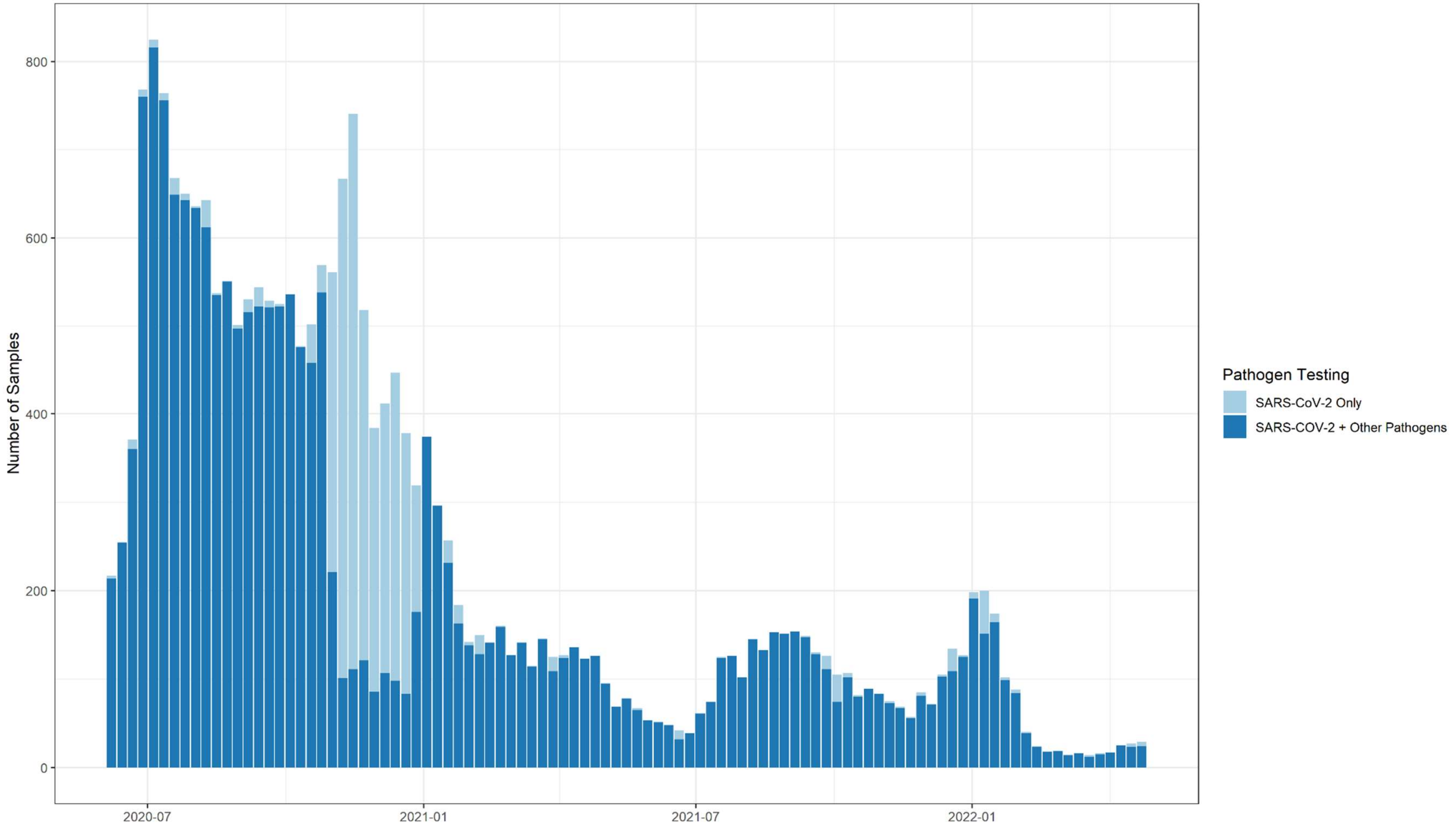

**eTable 1.** Comparison of participants who enrolled in SCAN multiple times versus participants who enrolled only once. For participants who enrolled multiple times, table data are based on the single observation included in the analysis.

| | Single Enrollment (N=17829) | Multiple Enrollments (N=5449) |
|---|---|---|
| **Number of enrollments** | | |
| Median [Min, Max] | 1.0 [1.0, 1.0] | 3.1[2.00, 25.0] |
| **Age** | | |
| Median [Min, Max] | 34.3 [0, 85.0] | 34.8 [0.3, 85.0] |
| **Age Group (years) N (%)** | | |
| 12-50 | 11858 (66.5%) | 4088 (75.0%) |
| <12 | 2541 (14.3%) | 570 (10.5%) |
| ≥50 | 3430 (19.2%) | 791 (14.5%) |
| **Sex** | | |
| Female | 10366 (58.1%) | 3388 (62.2%) |
| Male | 7463 (41.9%) | 2061 (37.8%) |
| **Race/ethnicity N (%)** | | |
| White | 11055 (62.0%) | 3438 (63.1%) |
| Asian | 2977 (16.7%) | 988 (18.1%) |
| Black | 515 (2.9%) | 136 (2.5%) |
| Hispanic | 1726 (9.7%) | 437 (8.0%) |
| Other | 1556 (8.7%) | 450 (8.3%) |
| **Region** | | |
| North | 12351 (69.3%) | 3629 (66.6%) |
| South | 5478 (30.7%) | 1820 (33.4%) |
| **Social & Economic Risk Index N (%)** | | |
| Low | 7040 (39.5%) | 2036 (37.4%) |
| Moderate | 5908 (33.1%) | 1839 (33.7%) |
| High | 4881 (27.4%) | 1574 (28.9%) |
| **Work Location N (%)** | | |
| Home | 5961 (33.4%) | 1963 (36.0%) |
| Home & onsite | 2083 (11.7%) | 812 (14.9%) |
| Onsite | 3147 (17.7%) | 1060 (19.5%) |
| Not employed | 6638 (37.2%) | 1614 (29.6%) |
| **Attend/work at school N (%)** | | |
| No | 15462 (86.7%) | 4622 (84.8%) |
| Yes | 2367 (13.3%) | 827 (15.2%) |

**eTable 2.** Comparison of the odds ratios and selected variables under different regression approaches: forward/backward stepwise regression using AIC, LASSO regression, and Bayesian Model Averaging.

| Variable | Stepwise AIC | LASSO (lambda min) | LASSO (lambda 1se) | BMA |
|---|---|---|---|---|
| <12 yrs | 1.17 | 1.12 | | |
| 50+ yrs | 1.22 | 1.17 | | |
| Male | | 1.04 | | |
| Asian | 1.05 | | | 1.06 |
| Black | 2.03 | 1.95 | 1.26 | 2.24 |
| Hispanic | 2.12 | 2.03 | 1.47 | 2.19 |
| Other Race | 1.31 | 1.25 | | 1.35 |
| South | 1.56 | 1.58 | 1.56 | 2.00 |
| Moderate | 1.25 | 1.18 | | |
| High | 1.55 | 1.49 | 1.19 | |
| 5+ ppl | 1.42 | 1.40 | 1.14 | 1.43 |
| Apartment | 1.16 | 1.13 | | |
| Other Housing | 1.42 | 1.36 | | |
| Confirmed Contact | 4.25 | 4.15 | 3.32 | 4.31 |
| Suspected Contact | 1.23 | 1.19 | | 1.25 |
| Home & Onsite | 0.76 | 0.78 | | 0.84 |
| Onsite | 1.03 | | | 1.05 |
| Not Employed | 1.19 | 1.17 | | 1.19 |
| School Attendance | 0.64 | 0.68 | | 0.66 |
| International Travel | 2.04 | 1.91 | | 1.23 |
| Domestic Travel | 1.52 | 1.44 | | 1.47 |
| Chills/Shivering | 1.68 | 1.67 | 1.50 | 1.77 |
| Cough | 2.39 | 2.33 | 1.92 | 2.43 |
| Runny/Stuffy Nose | 1.36 | 1.33 | | 1.33 |
| Sore Throat | 0.82 | 0.86 | | |
| Muscle/Body Aches | 1.47 | 1.47 | 1.17 | 1.54 |
| Fatigue | | 0.93 | | |
| Fever | 2.38 | 2.34 | 1.95 | 2.47 |
| Sweats | 1.17 | 1.15 | | |
| Diarrhea | 0.64 | 0.66 | | 0.66 |
| Trouble Breathing | 0.58 | 0.61 | | 0.58 |
| Nausea/Vomiting | 0.59 | 0.62 | | 0.61 |
| Rash | 0.42 | 0.47 | | 0.78 |
| Loss of Smell/Taste | 3.69 | 3.61 | 2.78 | 3.73 |
| Eye Pain | 1.59 | 1.55 | | 1.34 |
| Ear Pain/Discharge | | 0.97 | | |
| Headaches | | 1.01 | | |
| 3+ Symptoms | 1.21 | 1.21 | | |
| Prior Positive >90days | 0.38 | 0.41 | | 0.39 |
| No Prior Test | 1.24 | 1.20 | | 1.26 |
| Fully >6m | 1.03 | 1.07 | | 0.98 |
| Fully <6m | 0.73 | 0.79 | | 0.87 |
| Boosted | 0.61 | 0.66 | | 0.82 |
| Care Sought | 0.80 | 0.85 | | |
| Intermediate | 1.23 | 1.16 | | 1.16 |
| Omicron | 2.34 | 2.19 | 1.64 | 2.01 |
| Incidence | 1.71 | 1.69 | 1.52 | 1.72 |

**eFigure 3.** Cross tabulations between demographic variables. Reading from left to right, the numbers represent the proportion of each level from the top variable (x-axis) within the levels of the variables on the right (y-axis). For example, among children <12 years, .60 (60%) are White, .12 (12%) are Asian, etc. The shading represents the % positive for SARS-CoV-2 in that group. The highest % positive (~30%) is among Black participants living in households with ≥5 people.

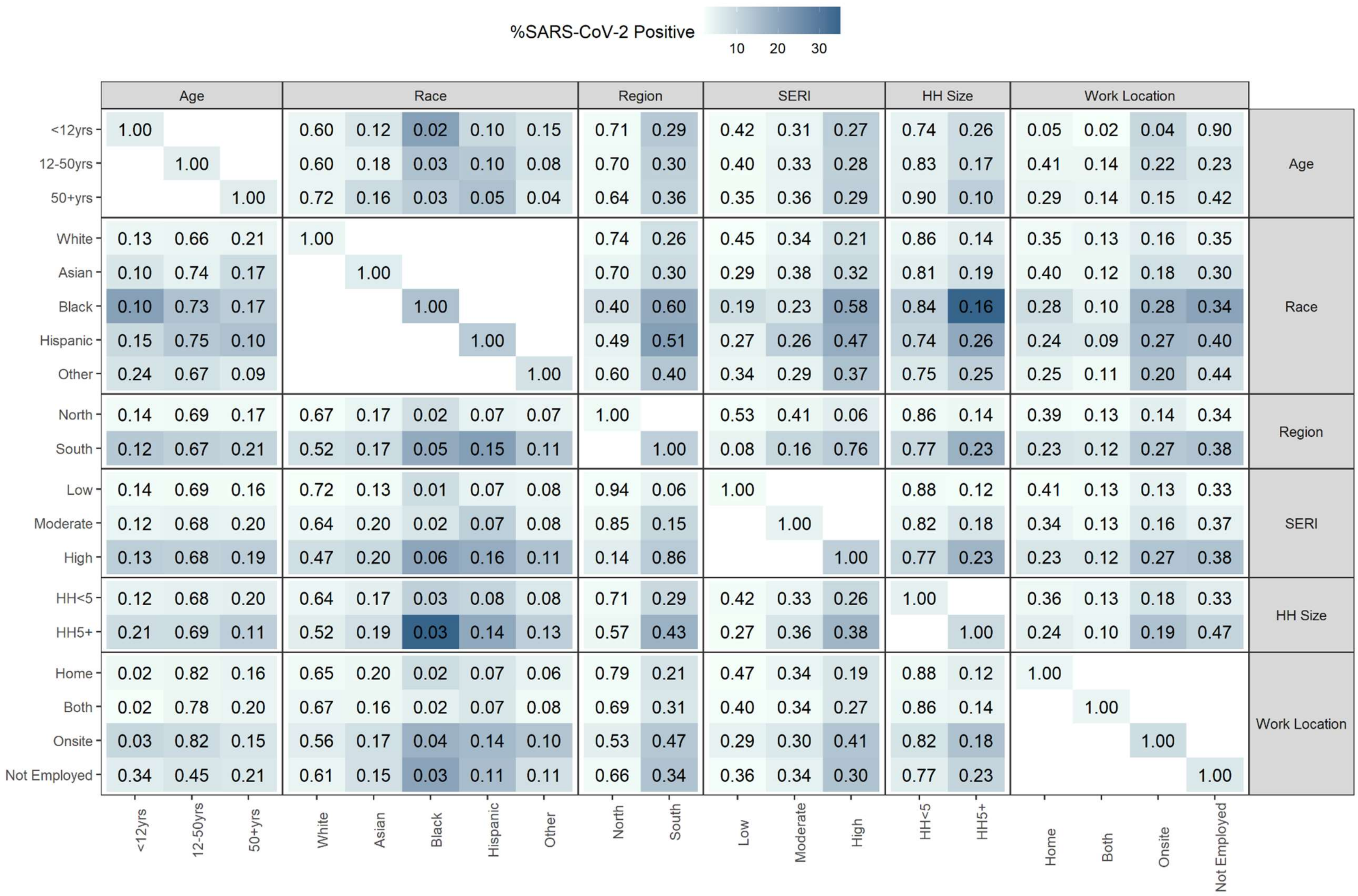


| | Age: <12yrs | Age: 12-50yrs | Age: 50+yrs | Race: White | Race: Asian | Race: Black | Race: Hispanic | Race: Other | Region: North | Region: South | SERI: Low | SERI: Moderate | SERI: High | HH Size: HH<5 | HH Size: HH5+ | Work Location: Home | Work Location: Both | Work Location: Onsite | Work Location: Not Employed |
|---|---|---|---|---|---|---|---|---|---|---|---|---|---|---|---|---|---|---|---|
| Age: <12yrs | 1.00 | | | 0.60 | 0.12 | 0.02 | 0.10 | 0.15 | 0.71 | 0.29 | 0.42 | 0.31 | 0.27 | 0.74 | 0.26 | 0.05 | 0.02 | 0.04 | 0.90 |
| Age: 12-50yrs | | 1.00 | | 0.60 | 0.18 | 0.03 | 0.10 | 0.08 | 0.70 | 0.30 | 0.40 | 0.33 | 0.28 | 0.83 | 0.17 | 0.41 | 0.14 | 0.22 | 0.23 |
| Age: 50+yrs | | | 1.00 | 0.72 | 0.16 | 0.03 | 0.05 | 0.04 | 0.64 | 0.36 | 0.35 | 0.36 | 0.29 | 0.90 | 0.10 | 0.29 | 0.14 | 0.15 | 0.42 |
| Race: White | 0.13 | 0.66 | 0.21 | 1.00 | | | | | 0.74 | 0.26 | 0.45 | 0.34 | 0.21 | 0.86 | 0.14 | 0.35 | 0.13 | 0.16 | 0.35 |
| Race: Asian | 0.10 | 0.74 | 0.17 | | 1.00 | | | | 0.70 | 0.30 | 0.29 | 0.38 | 0.32 | 0.81 | 0.19 | 0.40 | 0.12 | 0.18 | 0.30 |
| Race: Black | 0.10 | 0.73 | 0.17 | | | 1.00 | | | 0.40 | 0.60 | 0.19 | 0.23 | 0.58 | 0.84 | 0.16 | 0.28 | 0.10 | 0.28 | 0.34 |
| Race: Hispanic | 0.15 | 0.75 | 0.10 | | | | 1.00 | | 0.49 | 0.51 | 0.27 | 0.26 | 0.47 | 0.74 | 0.26 | 0.24 | 0.09 | 0.27 | 0.40 |
| Race: Other | 0.24 | 0.67 | 0.09 | | | | | 1.00 | 0.60 | 0.40 | 0.34 | 0.29 | 0.37 | 0.75 | 0.25 | 0.25 | 0.11 | 0.20 | 0.44 |
| Region: North | 0.14 | 0.69 | 0.17 | 0.67 | 0.17 | 0.02 | 0.07 | 0.07 | 1.00 | | 0.53 | 0.41 | 0.06 | 0.86 | 0.14 | 0.39 | 0.13 | 0.14 | 0.34 |
| Region: South | 0.12 | 0.67 | 0.21 | 0.52 | 0.17 | 0.05 | 0.15 | 0.11 | | 1.00 | 0.08 | 0.16 | 0.76 | 0.77 | 0.23 | 0.23 | 0.12 | 0.27 | 0.38 |
| SERI: Low | 0.14 | 0.69 | 0.16 | 0.72 | 0.13 | 0.01 | 0.07 | 0.08 | 0.94 | 0.06 | 1.00 | | | 0.88 | 0.12 | 0.41 | 0.13 | 0.13 | 0.33 |
| SERI: Moderate | 0.12 | 0.68 | 0.20 | 0.64 | 0.20 | 0.02 | 0.07 | 0.08 | 0.85 | 0.15 | | 1.00 | | 0.82 | 0.18 | 0.34 | 0.13 | 0.16 | 0.37 |
| SERI: High | 0.13 | 0.68 | 0.19 | 0.47 | 0.20 | 0.06 | 0.16 | 0.11 | 0.14 | 0.86 | | | 1.00 | 0.77 | 0.23 | 0.23 | 0.12 | 0.27 | 0.38 |
| HH Size: HH<5 | 0.12 | 0.68 | 0.20 | 0.64 | 0.17 | 0.03 | 0.08 | 0.08 | 0.71 | 0.29 | 0.42 | 0.33 | 0.26 | 1.00 | | 0.36 | 0.13 | 0.18 | 0.33 |
| HH Size: HH5+ | 0.21 | 0.69 | 0.11 | 0.52 | 0.19 | 0.03 | 0.14 | 0.13 | 0.57 | 0.43 | 0.27 | 0.36 | 0.38 | | 1.00 | 0.24 | 0.10 | 0.19 | 0.47 |
| Work Location: Home | 0.02 | 0.82 | 0.16 | 0.65 | 0.20 | 0.02 | 0.07 | 0.06 | 0.79 | 0.21 | 0.47 | 0.34 | 0.19 | 0.88 | 0.12 | 1.00 | | | |
| Work Location: Both | 0.02 | 0.78 | 0.20 | 0.67 | 0.16 | 0.02 | 0.07 | 0.08 | 0.69 | 0.31 | 0.40 | 0.34 | 0.27 | 0.86 | 0.14 | | 1.00 | | |
| Work Location: Onsite | 0.03 | 0.82 | 0.15 | 0.56 | 0.17 | 0.04 | 0.14 | 0.10 | 0.53 | 0.47 | 0.29 | 0.30 | 0.41 | 0.82 | 0.18 | | | 1.00 | |
| Work Location: Not Employed | 0.34 | 0.45 | 0.21 | 0.61 | 0.15 | 0.03 | 0.11 | 0.11 | 0.66 | 0.34 | 0.36 | 0.34 | 0.30 | 0.77 | 0.23 | | | | 1.00 |

**eTable 3.** Unadjusted and Adjusted Odds Ratios for SARS-CoV-2 test positivity, without interactions (*p<0.05; **p<0.001; ***p<0.0001).

| Characteristic | Univariable OR (95% CI) | Multivariable (No interactions) aOR (95% CI) |
|---|---|---|
| **Age** | | |
| 12 – 50 years (Ref) | 1.0 | 1.0 |
| <12 years | 1.0 (0.8- 1.2) | 1.2 (0.9- 1.5) |
| 50+ years | 1.0 (0.9- 1.2) | 1.2 (1.0- 1.5)* |
| **Sex** | | |
| Female (Ref) | 1.0 | 1.0 |
| Male | 1.0 (0.9- 1.2) | 1.0 (0.9- 1.2) |
| **Race/ethnicity** | | |
| White (ref) | 1.0 | 1.0 |
| Asian | 1.5 (1.2- 1.7)*** | 1.1 (0.9- 1.3) |
| Black | 3.8 (2.9- 4.8)*** | 2.0 (1.5- 2.7)*** |
| Hispanic | 3.3 (2.8- 3.8)*** | 2.1 (1.7- 2.6)*** |
| Other Race | 2.0 (1.6- 2.4)*** | 1.3 (1.1- 1.6)* |
| **Region** | | |
| North (Ref) | 1.0 | 1.0 |
| South | 3.8 (3.4- 4.3)*** | 1.6 (1.3- 1.9)*** |
| **SERI** | | |
| Low (Ref) | 1.0 | 1.0 |
| Moderate | 1.8 (1.5- 2.1)*** | 1.2 (1.0- 1.5)* |
| High | 4.6 (3.9- 5.3)*** | 1.6 (1.2- 2.0)** |
| **Household Size** | | |
| <5 people (Ref) | 1.0 | 1.0 |
| ≥5 people | 1.9 (1.7- 2.2)*** | 1.4 (1.2- 1.7)*** |
| **Housing Type** | | |
| House (Ref) | 1.0 | 1.0 |
| Apartment | 1.5 (1.3- 1.7)*** | 1.2 (1.0- 1.4) |
| Other Housing | 2.7 (1.8- 4.0)*** | 1.4 (0.9- 2.2) |
| **Contact with SARS-CoV-2 Case** | | |
| No Contact (Ref) | 1.0 | 1.0 |
| Confirmed Contact | 7.5 (6.6- 8.6)*** | 4.3 (3.7- 5.0)*** |
| Suspected/Possible Contact | 1.8 (1.5- 2.2)*** | 1.2 (1.0- 1.5)* |
| **Work Location** | | |
| Home (Ref) | 1.0 | 1.0 |
| Home & Onsite | 1.0 (0.8- 1.3) | 0.8 (0.6- 1.0)* |
| Onsite | 2.5 (2.1- 2.9)*** | 1.0 (0.8- 1.3) |
| Not Employed | 1.8 (1.6- 2.1)*** | 1.2 (1.0- 1.4) |
| **Binary Variables (Ref = No)** | 1.0 | 1.0 |
| Attend/Work at School | 0.8 (0.7- 1.0)* | 0.6 (0.5- 0.8)*** |
| International Travel | 1.5 (1.0- 2.1)* | 2.0 (1.3- 3.1)* |
| Domestic Travel | 0.9 (0.8- 1.1) | 1.5 (1.3- 1.8)*** |
| Clinical Care Sought | 1.8 (1.5- 2.1)*** | 0.8 (0.6- 1.0) |
| Chills/Shivering | 4.1 (3.6- 4.6)*** | 1.7 (1.4- 2.0)*** |

| Characteristic | Univariable OR (95% CI) | Multivariable (No interactions) aOR (95% CI) |
|---|---|---|
| Headaches | 2.1 (1.8- 2.3)*** | |
| Cough | 3.9 (3.4- 4.4)*** | 2.4 (2.0- 2.8)*** |
| Ear Pain/Discharge | 1.9 (1.5- 2.3)*** | |
| Runny/Stuffy Nose | 2.0 (1.8- 2.2)*** | 1.4 (1.2- 1.6)** |
| Sore Throat | 1.5 (1.4- 1.7)*** | 0.8 (0.7- 1.0)* |
| Muscle/Body Aches | 2.6 (2.3- 2.9)*** | 1.5 (1.2- 1.7)*** |
| Fatigue | 1.6 (1.4- 1.8)*** | |
| Fever | 4.0 (3.5- 4.5)*** | 2.4 (2.0- 2.8)*** |
| Sweats | 3.1 (2.7- 3.6)*** | 1.2 (1.0- 1.4) |
| Diarrhea | 1.0 (0.8- 1.1) | 0.6 (0.5- 0.8)*** |
| Trouble Breathing | 1.5 (1.2- 1.8)*** | 0.6 (0.5- 0.7)*** |
| Nausea/Vomiting | 1.3 (1.1- 1.5)* | 0.6 (0.5- 0.7)*** |
| Rash | 0.8 (0.5- 1.3) | 0.4 (0.2- 0.8)* |
| Loss of Smell/Taste | 5.7 (4.9- 6.7)*** | 3.7 (3.0- 4.5)*** |
| Eye Pain | 2.7 (2.3- 3.3)*** | 1.6 (1.2- 2.0)** |
| >3 Symptoms | 3.2 (2.9- 3.7)*** | 1.2 (1.0- 1.5)* |
| **Test History** | | |
| No Prior Positive (Ref) | 1.0 | 1.0 |
| Prior Positive (>90days) | 1.3 (0.8- 2.0) | 0.4 (0.2- 0.6)+* |
| No Prior Test | 0.7 (0.7- 0.8)*** | 1.2 (1.1- 1.4)* |
| **Vaccination Status** | | |
| Not Fully Vaccinated (Ref) | 1.0 | 1.0 |
| Fully >6m | 5.0 (4.1- 6.1)*** | 1.0 (0.8- 1.4) |
| Fully <6m | 1.7 (1.4- 2.0)*** | 0.7 (0.6- 1.0)* |
| Boosted | 4.3 (3.4- 5.3)*** | 0.6 (0.4- 0.9)* |
| **Time Period** | | |
| Early (Ref) | 1.0 | 1.0 |
| Intermediate | 1.7 (1.4- 1.9)*** | 1.2 (1.0- 1.5)* |
| Omicron | 10.2 (8.8-11.7)*** | 2.3 (1.7- 3.3)*** |
| **SARS-CoV-2 Incidence Indicator** | 2.5 (2.4- 2.7)*** | 1.7 (1.5- 1.9)*** |

**eFigure 4.** Sensitivity analysis for risk factors and symptoms associated with SARS-CoV-2 positivity. The first column displays the odds ratios from the final (core) model (without interactions), ordered from greatest to least. The middle column displays the odds ratios when the sample is restricted to pan-negative controls. The third column displays the odds ratios when the sample is restricted to participants reporting at least one symptom.

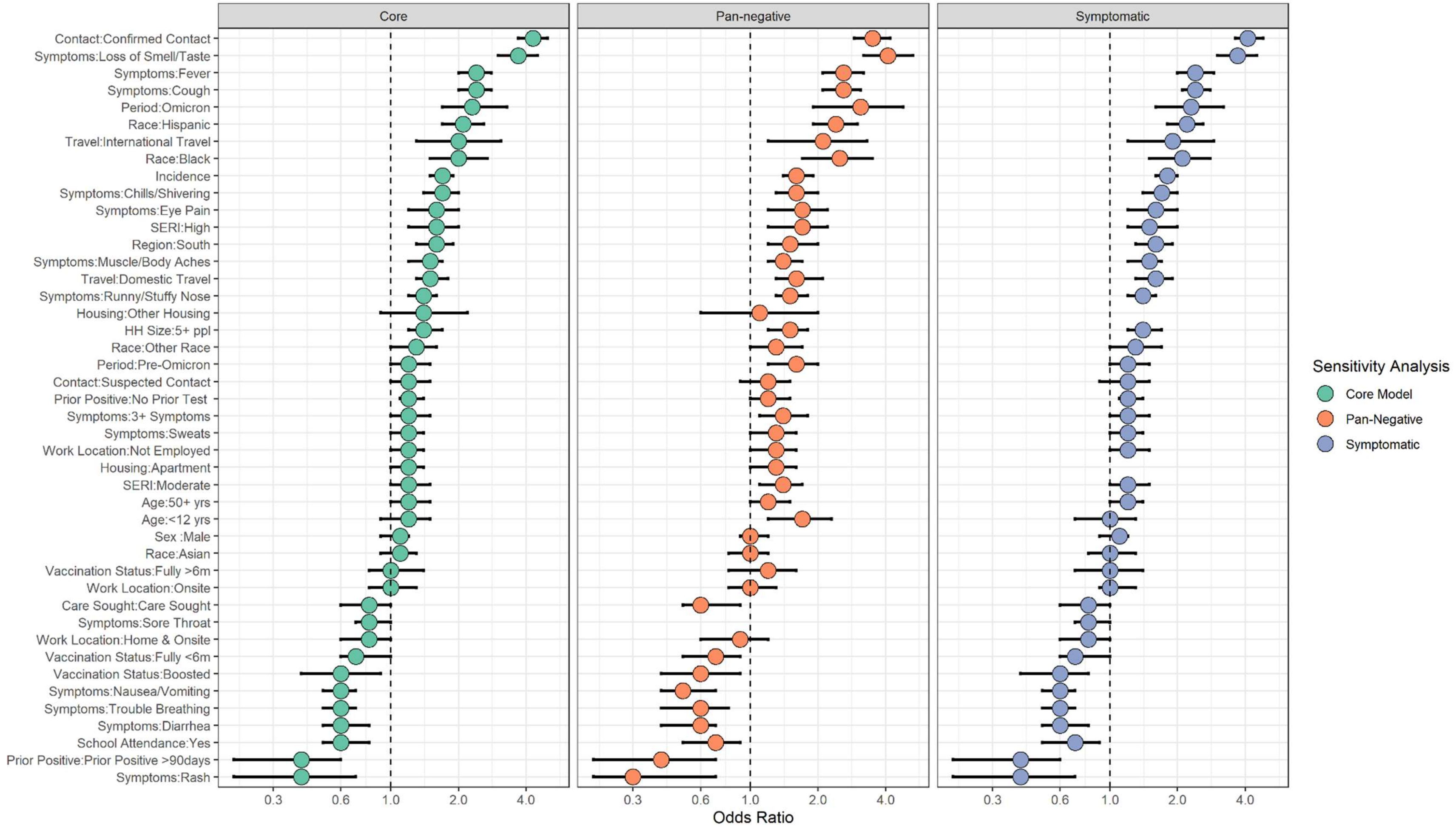

**eTable 4.** Unadjusted and Adjusted Odds Ratios for the Rhinovirus test positivity, without interactions (*p<0.05; **p<0.001; ***p<0.0001).

| variable | Univariable | Multivariable (No interactions) |
|---|---|---|
| **Age** | | |
| 12 – 50 years (Ref) | 1.0 | 1.0 |
| <12 yrs | 5.3 (4.8-5.8)*** | 4.0 (3.5-4.6)*** |
| 50+ yrs | 0.4 (0.3-0.4)*** | 0.4 (0.3-0.5)*** |
| **Sex** | | |
| Female (Ref) | 1.0 | 1.0 |
| Male | 1.1 (1.0-1.2) | 1.0 (0.9-1.1) |
| **Race/ethnicity** | | |
| White (ref) | 1.0 | 1.0 |
| Asian | 0.8 (0.7-0.9)*** | 0.9 (0.8-1.0)* |
| Black | 1.0 (0.8-1.3) | 1.4 (1.0-1.9)* |
| Hispanic | 1.2 (1.1-1.4)* | 1.1 (0.9-1.3) |
| Other Race | 1.3 (1.1-1.5)*** | 1.0 (0.8-1.1) |
| **Region** | | |
| North (Ref) | 1.0 | 1.0 |
| South | 1.2 (1.1-1.4)*** | 1.2 (1.0-1.5)* |
| **SERI** | | |
| Low (Ref) | 1.0 | 1.0 |
| Moderate | 1.1 (1.0-1.2) | 1.0 (0.9-1.1) |
| High | 1.2 (1.1-1.4)*** | 1.0 (0.8-1.2) |
| **Household Size** | | |
| <5 people (Ref) | 1.0 | 1.0 |
| 5+ ppl | 1.5 (1.3-1.6)*** | 1.1 (1.1-1.3)* |
| **Housing Type** | | |
| House (Ref) | 1.0 | 1.0 |
| Apartment | 0.6 (0.5-0.7)*** | 0.6 (0.6-0.7)*** |
| Other Housing | 0.3 (0.2-0.6)** | 0.4 (0.2-0.8)* |
| **Work Location** | | |
| Home (Ref) | 1.0 | 1.0 |
| Home & Onsite | 1.0(0.9-1.2) | |
| Onsite | 1.1 (1.0-1.3) | |
| Not Employed | 2.1 (1.9-2.3)*** | |
| **Binary Variables (Ref = No)** | 1.0 | 1.0 |
| School Attendance | 4.3 (3.9-4.7)*** | 2.0 (1.8-2.3)*** |
| International Travel | 0.7 (0.5-1.0)* | |
| Domestic Travel | 0.8 (0.8-1.0)** | |
| Clinical Care Sought | 1.1 (0.9-1.3) | |
| Chills/Shivering | 0.9 (0.8-1.1) | 0.7 (0.6-0.9)** |
| Headaches | 0.8 (0.7-0.8)*** | 0.8 (0.7-0.9)** |
| Cough | 2.9 (2.6-3.1)*** | 2.0 (1.8-2.2)*** |
| Ear Pain/Discharge | 1.4 (1.2-1.7)*** | 1.2 (1.0-1.5) |
| Runny/Stuffy Nose | 6.4 (5.7-7.1)*** | 4.6 (4.1-5.2)*** |
| Sore Throat | 2.3 (2.1-2.5)*** | 2.1 (1.9-2.3)*** |

| variable | Univariable | Multivariable (No interactions) |
|---|---|---|
| Muscle/Body Aches | 0.7 (0.6-0.8)*** | 0.9 (0.8-1.0)* |
| Fatigue | 1.1 (1.0-1.2) | |
| Fever | 1.9 (1.7-2.1)*** | 1.5 (1.3-1.7)*** |
| Sweats | 0.9(0.8-1.1) | 0.8 (0.7-1.0)* |
| Diarrhea | 0.6 (0.5-0.7)*** | 0.7 (0.6-0.8)*** |
| Trouble Breathing | 1.0 (0.8-1.1) | 0.8 (0.7-0.9)* |
| Nausea/Vomiting | 0.9 (0.8-1.0)* | 0.8 (0.7-1.0)* |
| Rash | 1.0 (0.7-1.3) | |
| Loss of Smell/Taste | 2.0 (1.6-2.3)*** | 2.0 (1.6-2.4)*** |
| Eye Pain | 0.9 (0.7-1.1) | 0.8 (0.6-1.1) |
| >3 Symptoms | 1.7(1.6-1.8)*** | |
| **Time Period** | | |
| Early (Ref) | 1.0 | 1.0 |
| Intermediate | 3.2 (3.0-3.5)*** | 1.3 (1.1-1.4)** |
| Omicron | 1.3(1.0-1.5)* | 1.1(0.8-1.3) |
| **Rhinovirus Incidence** | 1.1 (1.1-1.1)*** | 1.1 (1.1-1.1)*** |

**eFigure 5.** Sensitivity analysis for risk factors and symptoms associated with rhinovirus positivity. The first column displays the odds ratios from the final (core) model (without interactions), ordered from greatest to least. The middle column displays the odds ratios when the sample is restricted to pan-negative controls. The third column displays the odds ratios when the sample is restricted to participants reporting at least one symptom.

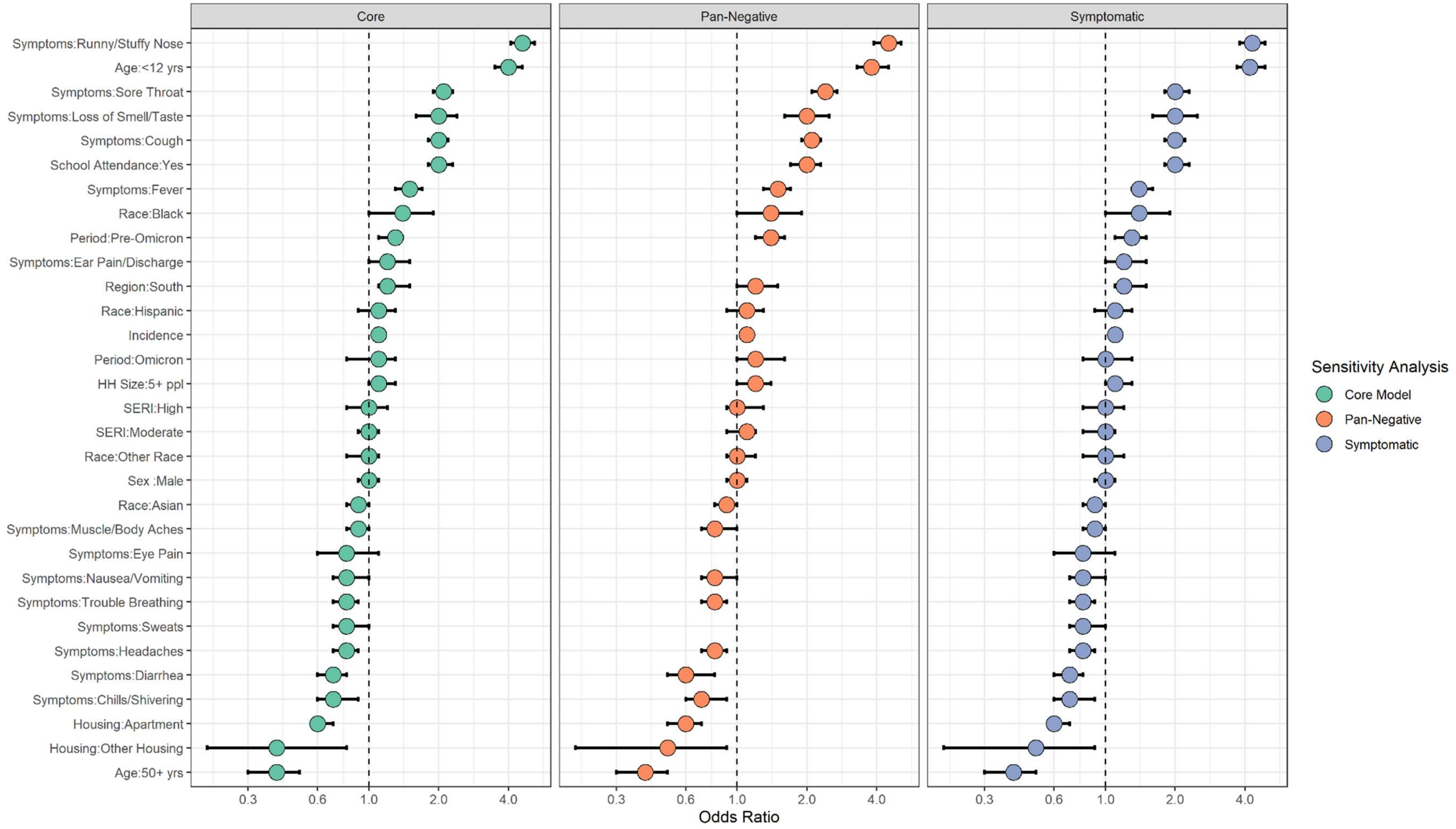

**eFigure 6.** Adjusted odds ratios for rhinovirus test positivity by time-period, for variables with significant interactions with time-period.

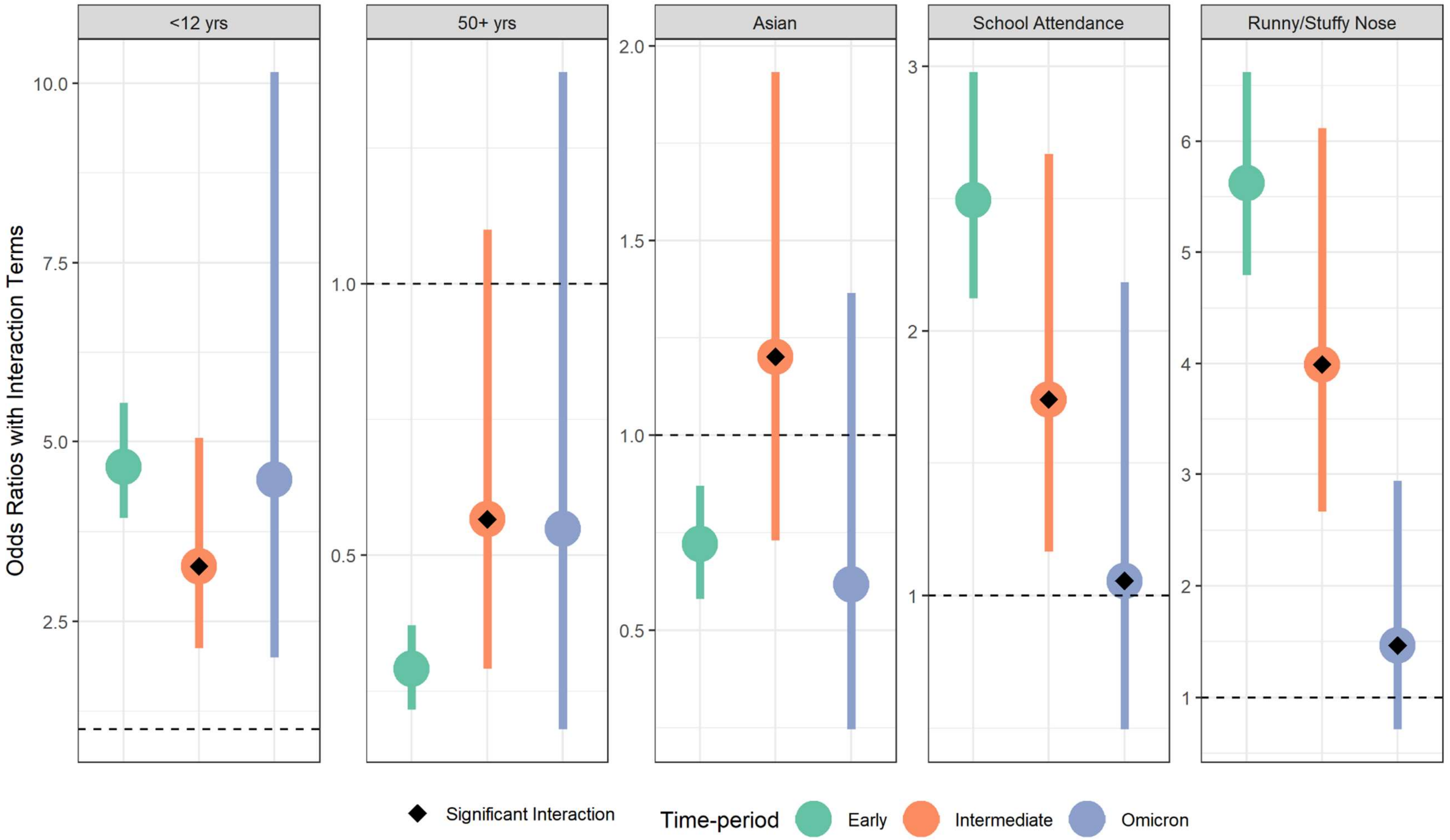

**eFigure 7.** Cross-correlation between rhinovirus and SARS-CoV-2 percent positive in SCAN data.

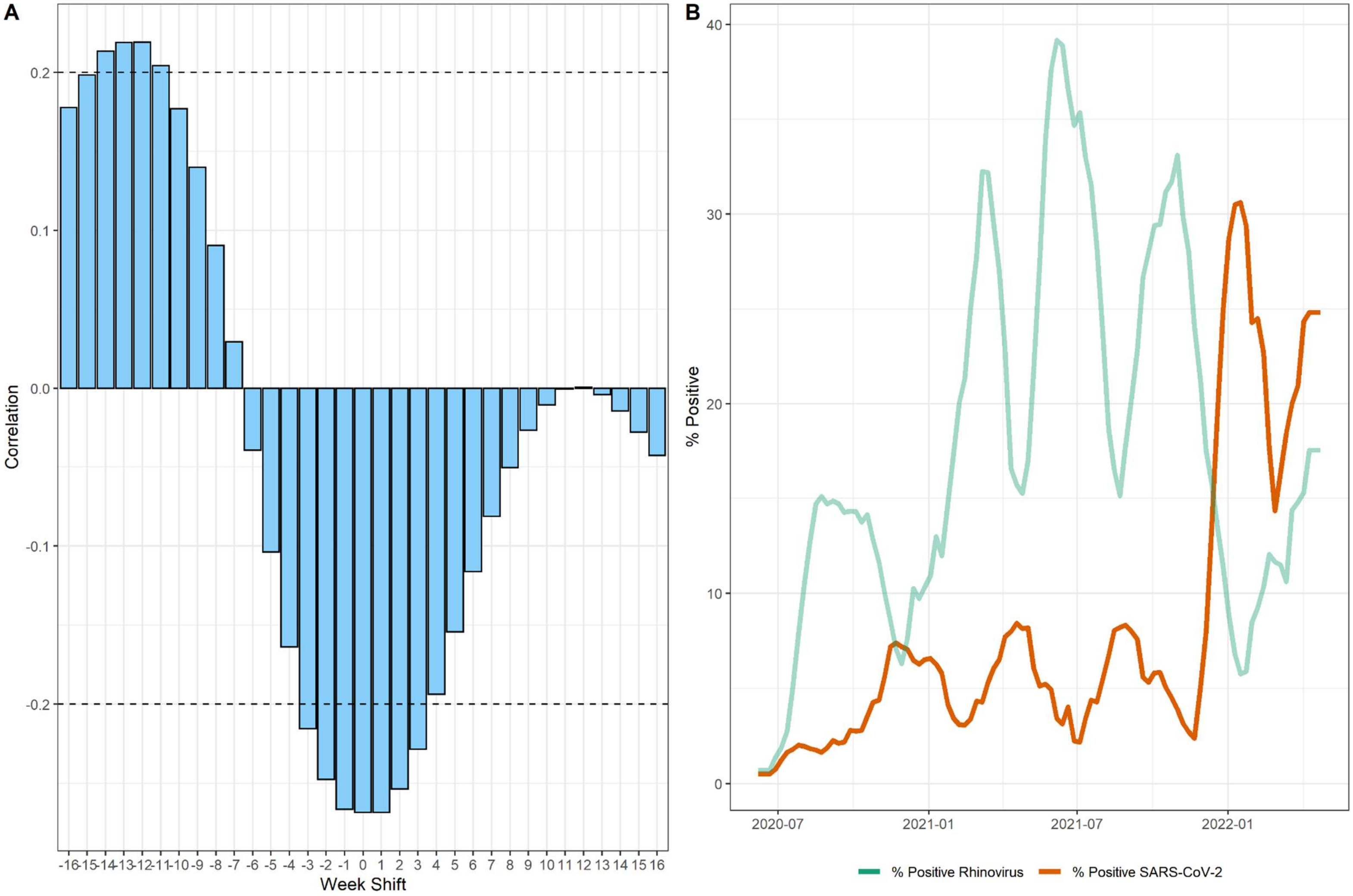

STROBE Statement—checklist of items that should be included in reports of observational studies

| | Item No | Recommendation | Page No |
|---|---|---|---|
| **Title and abstract** | 1 | (*a*) Indicate the study's design with a commonly used term in the title or the abstract | 1 |
| | | (*b*) Provide in the abstract an informative and balanced summary of what was done and what was found | 2-3 |
| **Introduction** | | | |
| Background/rationale | 2 | Explain the scientific background and rationale for the investigation being reported | 4 |
| Objectives | 3 | State specific objectives, including any prespecified hypotheses | 4 |
| **Methods** | | | |
| Study design | 4 | Present key elements of study design early in the paper | 5 |
| Setting | 5 | Describe the setting, locations, and relevant dates, including periods of recruitment, exposure, follow-up, and data collection | 5 |
| Participants | 6 | (*a*) *Cohort study*—Give the eligibility criteria, and the sources and methods of selection of participants. Describe methods of follow-up<br>*Case-control study*—Give the eligibility criteria, and the sources and methods of case ascertainment and control selection. Give the rationale for the choice of cases and controls<br>*Cross-sectional study*—Give the eligibility criteria, and the sources and methods of selection of participants | 5, supplement 2-3 |
| | | (*b*) *Cohort study*—For matched studies, give matching criteria and number of exposed and unexposed<br>*Case-control study*—For matched studies, give matching criteria and the number of controls per case | 5 |
| Variables | 7 | Clearly define all outcomes, exposures, predictors, potential confounders, and effect modifiers. Give diagnostic criteria, if applicable | 5-6, supplement 4-6 |
| Data sources/ measurement | 8* | For each variable of interest, give sources of data and details of methods of assessment (measurement). Describe comparability of assessment methods if there is more than one group | 5-6 |
| Bias | 9 | Describe any efforts to address potential sources of bias | Supplement 8 |
| Study size | 10 | Explain how the study size was arrived at | Supplement 19 |
| Quantitative variables | 11 | Explain how quantitative variables were handled in the analyses. If applicable, describe which groupings were chosen and why | Supplement 4-6 |
| Statistical methods | 12 | (*a*) Describe all statistical methods, including those used to control for confounding | 6 |
| | | (*b*) Describe any methods used to examine subgroups and interactions | 6 |
| | | (*c*) Explain how missing data were addressed | 5 |
| | | (*d*) *Cohort study*—If applicable, explain how loss to follow-up was addressed | Supplement 8 |

| | | |
|---|---|---|
| | *Case-control study*—If applicable, explain how matching of cases and controls was addressed<br>*Cross-sectional study*—If applicable, describe analytical methods taking account of sampling strategy | |
| | (e) Describe any sensitivity analyses | Supplement 7-8 |

Continued on next page

| | | | |
|---|---|---|---|
| **Results** | | | |
| Participants | 13* | (a) Report numbers of individuals at each stage of study—eg numbers potentially eligible, examined for eligibility, confirmed eligible, included in the study, completing follow-up, and analysed | 7 |
| | | (b) Give reasons for non-participation at each stage | |
| | | (c) Consider use of a flow diagram | Supplement 19 |
| Descriptive data | 14* | (a) Give characteristics of study participants (eg demographic, clinical, social) and information on exposures and potential confounders | 7, 20 |
| | | (b) Indicate number of participants with missing data for each variable of interest | |
| | | (c) *Cohort study*—Summarise follow-up time (eg, average and total amount) | |
| Outcome data | 15* | *Cohort study*—Report numbers of outcome events or summary measures over time | |
| | | *Case-control study*—Report numbers in each exposure category, or summary measures of exposure | |
| | | *Cross-sectional study*—Report numbers of outcome events or summary measures | *20* |
| Main results | 16 | (*a*) Give unadjusted estimates and, if applicable, confounder-adjusted estimates and their precision (eg, 95% confidence interval). Make clear which confounders were adjusted for and why they were included | 7-9, Supplement 24-25 |
| | | (*b*) Report category boundaries when continuous variables were categorized | |
| | | (*c*) If relevant, consider translating estimates of relative risk into absolute risk for a meaningful time period | |
| Other analyses | 17 | Report other analyses done—eg analyses of subgroups and interactions, and sensitivity analyses | 8 |
| **Discussion** | | | |
| Key results | 18 | Summarise key results with reference to study objectives | 9 |
| Limitations | 19 | Discuss limitations of the study, taking into account sources of potential bias or imprecision. Discuss both direction and magnitude of any potential bias | 12-13 |
| Interpretation | 20 | Give a cautious overall interpretation of results considering objectives, limitations, multiplicity of analyses, results from similar studies, and other relevant evidence | 13 |
| Generalisability | 21 | Discuss the generalisability (external validity) of the study results | 13 |
| **Other information** | | | |
| Funding | 22 | Give the source of funding and the role of the funders for the present study and, if applicable, for the original study on which the present article is based | 14 |

*Give information separately for cases and controls in case-control studies and, if applicable, for exposed and unexposed groups in cohort and cross-sectional studies.

**Note:** An Explanation and Elaboration article discusses each checklist item and gives methodological background and published examples of transparent reporting. The STROBE checklist is best used in conjunction with this article (freely available on the Web sites of PLoS Medicine at http://www.plosmedicine.org/, Annals of Internal Medicine at http://www.annals.org/, and Epidemiology at http://www.epidem.com/). Information on the STROBE Initiative is available at www.strobe-statement.org.
